# Factors associated with excess all-cause mortality in the first wave of COVID-19 pandemic in the UK: a time-series analysis using the Clinical Practice Research Datalink

**DOI:** 10.1101/2021.06.04.21258344

**Authors:** Helen Strongman, Helena Carreira, Bianca L De Stavola, Krishnan Bhaskaran, David A Leon

## Abstract

**Objectives:** Excess mortality captures the total effect of the COVID-19 pandemic on mortality and is not affected by mis-specification of cause of death. We aimed to describe how health and demographic factors have been associated with excess mortality during the pandemic.

**Design:** Time-series analysis.

**Setting:** UK primary care data from practices contributing to the Clinical Practice Research Datalink on July 31^st^ 2020.

**Participants:** We constructed a time-series dataset including 9,635,613 adults (≥40 years old) who were actively registered at the general practice during the study period.

**Main outcome measures:** We extracted weekly numbers of deaths between March 2015 and July 2020, stratified by individual-level factors. Excess mortality during wave 1 of the UK pandemic (5th March to 27th May 2020) compared to pre-pandemic was estimated using seasonally adjusted negative binomial regression models. Relative rates of death for a range of factors were estimated before and during wave 1 by including interaction terms.

**Results:** All-cause mortality increased by 43% (95% CI 40%-47%) during wave 1 compared with pre-pandemic. Changes to the relative rate of death associated with most socio-demographic and clinical characteristics were small during wave 1 compared with pre-pandemic. However, the mortality rate associated with dementia markedly increased (RR for dementia vs no dementia pre-pandemic: 3.5, 95% CI 3.4-3.5; RR during wave 1: 5.1, 4.87-5.28); a similar pattern was seen for learning disabilities (RR pre-pandemic: 3.6, 3.4-3.5; during wave 1: 4.8, 4.4-5.3), for Black or South Asian ethnicity compared to white, and for London compared to other regions.

**Conclusions:** The first UK COVID-19 wave appeared to amplify baseline mortality risk by a relatively constant factor for most population subgroups. However disproportionate increases in mortality were seen for those with dementia, learning disabilities, non-white ethnicity, or living in London.

**Summary box:** *What is already known on this topic:*

- All-cause mortality during the COVID-19 pandemic was higher than in previous years; this excess mortality was particularly pronounced among elderly people, males, people of non-white ethnicity, people of lower socio-economic status and people living in care-homes.
- Several other papers have studied a wider range of factors associated with mortality due to COVID-19 using cause-of-death data.
- There is little evidence on how all-cause mortality has changed in people with comorbidities.

*What this study adds:*

- Our study shows that during Wave 1 of the pandemic all cause death rates increased by a similar proportional degree for almost all population subgroups regardless of their health or socio-demographic circumstances; the exceptions were those with a diagnosis of dementia or learning disabilities and those of non-white ethnicity or living in London.
- This suggests that COVID-19 has dialled up the risk of death by a similar proportional degree for everyone except those exposed to a higher risk of infection.

## INTRODUCTION

Understanding how pre-existing health status and demographic factors affect the risk of infection and death from COVID-19 has been a critical clinical and public health priority since the start of the pandemic. This knowledge informs optimal care and treatment of those infected with SARS-CoV-2 as well as policies on shielding and vaccination. A number of studies of COVID-19 mortality in relation to health and demographic factors have highlighted factors such as age, deprivation, ethnicity and diabetes as particular risk factors for death from COVID-19[1–9]. Studies of the impact of health and demographic factors on mortality from conditions other than COVID-19 during the pandemic have been relatively uncommon. An analysis of over 17 million adults registered with primary care practices in England[10] found that most factors associated with COVID-19 death were similarly associated with non-COVID death. A key limitation of this study is reliance on cause-specific mortality data. In the UK, in the first months of the 2020 pandemic, it is very likely that attribution of deaths to COVID-19 was unreliable because of the novelty of the virus, uncertainty about its precise clinical manifestations and an absence of wide-spread testing for laboratory confirmation of infection. The problem of attribution of cause is further complicated by the probability that some deaths may have occurred as a result of indirect effects, such as reduced diagnosis and care for non-COVID conditions, plus the broader effect of voluntary and obligatory behavioural change such as social distancing.

One approach to circumventing the challenge of analysing cause-specific mortality has been to assess the impact of the pandemic on total excess deaths[11], a methodology that compares the number of deaths from any cause during the pandemic with the number expected in the same period based on mortality rates in earlier (pre-pandemic) years; this has been previously used to study the impact of seasonal influenza and acute exposures such as heat waves. Total excess mortality captures both the direct and indirect impacts of COVID-19 on mortality and is not affected by validity of cause of death. Excess mortality rates during the pandemic have been estimated for national and sub-national populations[12–15]. de Lusignan et al. compared mortality in patients registered in a sample of general practices during Wave 1 with external life table data from 2018 to show that excess mortality may be particularly raised in men, older individuals, black people, and those with certain comorbidities; key factors such as dementia were not studied[16]. There are no published analyses covering a wide range of health and demographic factors and comparing mortality rates pre-pandemic with those in the pandemic period within the same study population. Observational studies have therefore assumed that relative excess mortality in people with and without comorbidities was the same during and prior to the pandemic[16,17].

We examined excess mortality in the UK during Wave 1 of the COVID-19 pandemic according to an extensive range of morbidities (including dementia) and socio-demographic characteristics. To do this we used a large regularly updated primary care database covering 20% of the UK population.

## METHODS

### Study design and setting

We conducted a population-based time-series study using data prospectively collected from 5^th^ March 2015 to 31st July 2020 from the UK Clinical Practice Research Datalink (CPRD).

General practices have an important role in the UK’s National Health Services, with responsibility for primary care and specialist referrals for the vast majority of the UK population[18]. General Practitioners (GPs) record clinical diagnoses, hospital diagnoses that affect patients’ ongoing care, primary care prescribing, test and laboratory data, in specialised software systems. Clinical diagnoses are recorded using Read or Snomed codes. In our primary analyses, we used data from CPRD GOLD[19] and CPRD Aurum[20], which include anonymised data collected from two software systems. Individually, these databases are broadly representative of the UK population in terms of age and sex[19,20]. Together, they cover over 20% of the UK population, distributed across all UK countries and English regions.

CPRD primary care data may be linked to additional health and area-based deprivation data sets. Quintiles of the Carstairs Index and urban-rural data were added by CPRD through linkage using the practice postcode[21]. Linkages based on patient data are also available[22], but are restricted to English practices that have agreed to participate in the linkage programme, reducing geographic coverage and the sample size. In sensitivity analyses, we used the Office for National Statistics (ONS) mortality data (national death registration data), the ICD-10 coded clinical data from Hospital Episode Statistics Admitted Patient Care (HES APC)[23], and patient-level quintile of Carstairs Index.

### Study population and procedures

We constructed a time series dataset from a study population that included all adults (≥40 years old) actively registered during the study period in a general practice contributing to CPRD on 31st July 2020. Follow-up in the base population started at the latest of 5th March 2015, 40th birthday, and one year after registration with the practice. Follow-up ended at the earliest of the end of the study period (i.e. 31^st^ July 2020), the date the patient left the practice or died. Birth dates were estimated at 1st July of the year of birth as the day and month are not collected by CPRD. Date of death was identified from the CPRD-derived death date, which uses data recorded in general practices and has been shown to agree closely with the ONS registered death date[24].

Number of deaths and number at risk were counted for each week of the year from 5th March 2015 - 31st July 2020. Each week was classified as “pre-pandemic” (before 5th March 2020), during Wave 1 of the pandemic (5th March to 27th May 2020), and after Wave 1 (28th May to 31st July).

Demographic factors included 5-year age groups, sexes, deprivation quintiles, ethnicities and regions. Health factors included Body Mass Index (BMI) categories, smoking status and morbidities. Weekly numbers of deaths and numbers at risk were obtained separately for people with each health and demographic factor. For all morbidities except asthma and cancer, individuals were included in the exposed numbers at risk (of death) if they had a record of that morbidity prior to the start of the week. Asthma counts required a record in the last three years; for cancer, the first ever record was required to be in the past year. Weekly numbers of deaths and numbers at risk for people without each morbidity (the unexposed) were obtained by subtracting counts for each morbidity from the total study population counts. For BMI and smoking status, the most recent records prior to each week were used to define the category at the start of each week. For demographic factors other than age, categories were assigned at the start of follow-up. For descriptive purposes, counts were obtained for missing ethnicity, BMI and smoking status.

A complete list of the factors included in the analyses is given in table 1; full definitions are provided in the supplementary appendix (p22-24). Code lists for all study variables are available online (https://doi.org/10.17037/DATA.00002269).

**Table 1:** Number of person-weeks and observed, expected and excess number of deaths during Wave 1 (5th March to 27th May 2020)

| Stratifying factor | Person-weeks in millions (% weeks in base population) | Observed deaths (% deaths in study population) | Observed deaths per million | Expected deaths per million-person weeks (95% CI) | Excess deaths per million-person weeks (95% CI) |
| --- | --- | --- | --- | --- | --- |
| Study population | 88.1 (100.0) | 35,369 (100.0) | 401 | 272 (271-272) | 130 (129-130) |
| **Age group** |  |  |  |  |  |
| 40 to 49 | 23.7 (26.9) | 894 (2.5) | 38 | 30 (30-30) | 8 (8-8) |
| 50 to 59 | 24.4 (27.7) | 2,262 (6.4) | 93 | 66 (66-66) | 26 (26-27) |
| 60 to 69 | 18.1 (20.6) | 4,115 (11.6) | 227 | 165 (164-165) | 62 (62-63) |
| 70 to 79 | 13.9 (15.8) | 8,253 (23.3) | 593 | 411 (410-412) | 181 (180-182) |
| 80 plus | 7.9 (9.0) | 19,845 (56.1) | 2504 | 1,619 (1,614-1,624) | 885 (880-890) |
| **Sex** |  |  |  |  |  |
| male | 43.5 (49.3) | 17,907 (50.6) | 412 | 276 (275-277) | 136 (136-137) |
| female | 44.7 (50.7) | 17,462 (49.4) | 391 | 268 (267-268) | 123 (123-124) |
| **Deprivation** |  |  |  |  |  |
| 1 (least deprived) | 12.6 (14.3) | 4,502 (12.7) | 358 | 239 (239-240) | 119 (118-120) |
| 2 | 17.5 (19.8) | 6,764 (19.1) | 387 | 264 (264-265) | 123 (122-124) |
| 3 | 20.1 (22.8) | 8,186 (23.1) | 408 | 283 (282-284) | 125 (125-126) |
| 4 | 18.9 (21.4) | 7,922 (22.4) | 420 | 290 (290-291) | 129 (128-130) |
| 5 (most deprived) | 17.5 (19.8) | 7,416 (21.0) | 425 | 268 (267-269) | 157 (156-158) |
| **Urban Rural\$** | | | | | |
| Urban | 75.77 (86.0) | 30,639 (86.6) | 404 | 270 (270-271) | 134 (133-135) |
| Rural | 12.60 (14.3) | 4,865 (13.8) | 386 | 283 (282-284) | 103 (102-104) |
| **Ethnicity** |  |  |  |  |  |
| White | 57.4 (65.1) | 23,473 (66.4) | 409 | 282 (281-283) | 127 (127-128) |
| South Asian | 4.2 (4.7) | 1,057 (3.0) | 254 | 128 (127-129) | 126 (125-127) |
| Black | 2.7 (3.0) | 904 (2.6) | 339 | 138 (137-139) | 201 (200-203) |
| Other and mixed | 1.9 (2.2) | 369 (1.0) | 189 | 88 (87-89) | 102 (101-103) |
| Missing | 22.2 (25.2) | 9,701 (27.4) | 437 | 307 (306-308) | 130 (129-131) |
| **Region** |  |  |  |  |  |
| North East | 2.35 (2.7) | 1,194 (3.4) | 507 | 325 (323-327) | 183 (181-185) |
| North West | 13.30 (15.1) | 5,910 (16.7) | 444 | 299 (298-300) | 146 (145-147) |
| Yorkshire & the Humber | 2.4 (2.7) | 941 (2.7) | 401 | 296 (295-298) | 105 (103-107) |
| East Midlands & East of England | 4.5 (5.1) | 1,630.0 (4.6) | 365 | 250 (248-251) | 115 (114-116) |
| West Midlands | 11.9 (13.5) | 5,042 (14.3) | 425 | 283 (282-284) | 142 (141-143) |
| South West | 8.7 (9.9) | 3,207 (9.1) | 369 | 278 (277-279) | 91 (90-92) |
| South Central | 9.2 (10.4) | 3,451 (9.8) | 377 | 254 (253-255) | 123 (122-124) |
| London | 13.2 (15.0) | 4,858 (13.7) | 367 | 192 (192-193) | 175 (174-175) |
| South East Coast | 7.6 (8.6) | 2,964 (8.4) | 389 | 262 (261-263) | 128 (127-129) |
| Northern Ireland | 1.9 (2.2) | 714 (2.0) | 369 | 292 (290-294) | 77 (75-79) |
| Scotland | 7.8 (8.8) | 3,152 (8.9) | 405 | 309 (308-310) | 96 (95-97) |
| Wales | 5.4 (6.1) | 2,306 (6.5) | 429 | 327 (326-329) | 102 (100-104) |

| Stratifying factor | Person-weeks in millions (% weeks in base population) | Observed deaths (% deaths in study population) | Observed deaths per million | Expected deaths per million weeks (95% CI) | Excess deaths per million weeks (95% CI) |
| --- | --- | --- | --- | --- | --- |
| **Morbidity** |  |  |  |  |  |
| **Autoimmune condition** |  |  |  |  |  |
| Psoriasis | 4.2 (4.8) | 2,019.0 (5.7) | 479 | 324 (323-326) | 155 (153-156) |
| Rheumatoid arthritis | 1.3 (1.4) | 1,100.0 (3.1) | 872 | 588 (585-592) | 284 (280-287) |
| **Cardiovascular disease** |  |  |  |  |  |
| Cerebrovascular disease | 3.7 (4.2) | 7,159 (20.2) | 1930 | 1,274 (1,270-1,278) | 656 (652-660) |
| Venous thromboembolism | 2.6 (3.0) | 3,844 (10.9) | 1453 | 1,016 (1,012-1,020) | 437 (433-441) |
| Chronic heart disease | 7.5 (8.5) | 11,885 (33.6) | 1590 | 1,113 (1,110-1,117) | 477 (474-480) |
| Hypertension | 35.3 (40.1) | 25,893 (73.2) | 733 | 498 (497-499) | 235 (234-237) |
| **Chronic respiratory disease** |  |  |  |  |  |
| Asthma | 8.0 (9.1) | 3,367.0 (9.5) | 420 | 291 (290-293) | 129 (128-130) |
| Other | 5.2 (5.9) | 7,111.0 (20.1) | 1376 | 1,040 (1,036-1,043) | 336 (333-340) |
| **Neurological conditions associated with respiratory infection** |  |  |  |  |  |
| Dementia | 1.8 (2.0) | 9,937 (28.1) | 5618 | 2,929 (2,918-2,941) | 2,688 (2,677-2,699) |
| Learning disabilities | 0.4 (0.5) | 397 (1.1) | 994 | 433 (427-438) | 561 (556-567) |
| Other | 1.5 (1.7) | 2,314 (6.5) | 1531 | 971 (967-976) | 560 (555-564) |
| **Other morbidity** |  |  |  |  |  |
| Chronic kidney disease | 12.1 (13.7) | 20,255 (57.3) | 1677 | 1,094 (1,091-1,097) | 583 (580-586) |
| Cancer (diagnosed in last year) | 0.6 (0.7) | 2,740 (7.7) | 4275 | 3,659 (3,647-3,671) | 616 (603-628) |
| Diabetes | 9.8 (11.2) | 9,878 (27.9) | 1003 | 643 (641-645) | 361 (359-363) |
| Multimorbidity | 3.5 (4.0) | 6,780 (19.2) | 1929 | 1,398 (1,394-1,402) | 531 (526-535) |
| **Health Indicators** |  |  |  |  |  |
| **Body Mass Index** |  |  |  |  |  |
| <18.5 (Underweight) | 1.3 (1.4) | 3,144 (8.9) | 2465 | 1,830 (1,823-1,838) | 635 (627-643) |
| 18.5-<25 (Normal weight) | 26.7 (30.3) | 13,002 (36.8) | 487 | 338 (337-339) | 149 (148-150) |
| 25-<30 (Overweight) | 30.3 (34.4) | 9,592 (27.1) | 316 | 219 (218-219) | 98 (97-98) |
| 30-<35 (Obesity class I) | 15.2 (17.2) | 4,703 (13.3) | 310 | 195 (194-196) | 115 (114-115) |
| >=35 (Obesity class II plus) | 8.6 (9.8) | 2,868.0 (8.1) | 333 | 216 (215-217) | 117 (116-118) |
| Missing | 6.3 (7.1) | 2,166 (6.1) | 344 | 195 (194-196) | 149 (148-150) |
| **Smoking status** |  |  |  |  |  |
| Non-smoker | 28.1 (31.9) | 8,400 (23.7) | 299 | 187 (187-188) | 111 (111-112) |
| Current smoker | 9.7 (11.0) | 3,539 (10.0) | 367 | 305 (304-306) | 61 (60-62) |
| Ex-smoker | 48.3 (54.8) | 22,678 (64.1) | 470 | 318 (317-319) | 152 (151-152) |
| Missing | 2.3 (2.6) | 863 (2.4) | 377 | 200 (199-202) | 177 (175-178) |

Legend: \*Carstairs Index. This is not available in Northern Ireland. <sup>\$</sup>2 mixed Urban-Rural practices in Northern Ireland were reclassified as Urban. For time-updating stratifying factors (e.g. BMI and smoking status), the sums of person weeks in millions and observed deaths do not equal the study population. This is because periods of person time were counted from the first full week of follow-up. Short periods of person time from the date the stratifying factor changed to the start of the following week were not therefore counted.

### Statistical analysis

We fitted generalised linear models with a negative binomial error structure to the weekly counts of deaths, taking account of the numbers at risk in each week (the denominator sizes[25], by offsetting it after log-transformation). All models included Fourier terms (for creating a harmonic function of calendar week) to capture seasonality in death rates, and a quadratic function of calendar year (treated as a continuous variable) to capture any major underlying trends. We refer to this as the basic model.

#### Overall and Relative Excess mortality

The basic model was fitted using only the data from the pre-pandemic period, initially for the overall population and then restricted to each factor in turn (e.g. black ethnicity, overweight, dementia, diabetes), to predict the number of deaths to be expected during the pandemic period (during and after Wave 1). We visually checked the adequacy of our generalised linear models by comparing observed and expected deaths in the pre-pandemic period. We calculated excess deaths during Wave 1 of the pandemic by subtracting total predicted deaths during this period from total observed deaths. We estimated 95% confidence intervals by pooling the weekly standard errors for predicted deaths.[26]

#### Association between individual factors and mortality before and during Wave 1

We estimated relative rates of death (RR) for each health and demographic factor in separate models, allowing for an interaction with the time period (before vs during Wave 1 of the pandemic) and adjusting for age and sex differences. This was done by extending the basic model to include a binary pandemic indicator (1 for Wave 1, 0 otherwise), the binary/categorical variable representing the health or demographic factor grouping (e.g. diabetes, ethnicity, BMI), and terms capturing their interaction. Age (in 5-year age groups) and sex were also included in each model to adjust for differences in age and sex structure by health or socio-demographic status (e.g. people with or without diabetes, black ethnicity versus white ethnicity). Missing categories for ethnicity, BMI and smoking status were excluded in these primary analyses. We examined partial autocorrelation plots and found no evidence of residual autocorrelation in the final models[8].

#### Secondary and sensitivity analyses

In secondary analyses, we described differences between age groups and sex. For cancer, previous evidence shows an increased risk of poor COVID-19 outcomes in haematological cancer patients[1], and thus we also estimated relative rates of death separately for haematological and non-haematological cancer patients, all diagnosed in the last year. As people recently diagnosed with cancer may have been more likely to avoid infection than long-term cancer survivors, we further extended our analyses to include people who had cancer diagnosed >1 year and <5 years ago, and more than 5 years ago.

To examine the robustness of our findings we completed sensitivity analyses using additional linked data. The study population was restricted to individuals registered in English practices who were included in CPRD’s linkage programme[27]. The ONS mortality date was used in place of the CPRD derived death date, HES data were used in addition to primary care data to define ethnicity and morbidities (except asthma), and the patient postcode level Carstairs index was used to define deprivation. Additional sensitivity analyses compared findings using the CPRD GOLD and Aurum databases and the addition of a missing data category to the RR analyses for ethnicity, BMI and smoking status.

We used StataMP 16.0 for all analyses.

### Patient public involvement

Patients and the public were not involved in conceiving, designing or conducting this study and will not be consulted regarding the dissemination of study results.

## RESULTS

Our primary analysis was based on 9,635,613 individuals aged at least 40 with a minimum 1 year of follow-up and who were registered in 1,754 practices that contributed data to CPRD GOLD or Aurum on 31^st^ July 2020 (Figure 1). Median follow-up in the base population was 5.4 years (IQR 2.3-5.4) with individuals contributing 1989.8 million-person weeks of follow-up, 88.1 million-person weeks of which were during Wave 1. A total of 585,170 deaths were observed of which 35,369 were in Wave 1. The number of individuals contributing each week increased gradually over time (supplementary appendix p24).

**Figure 1:**
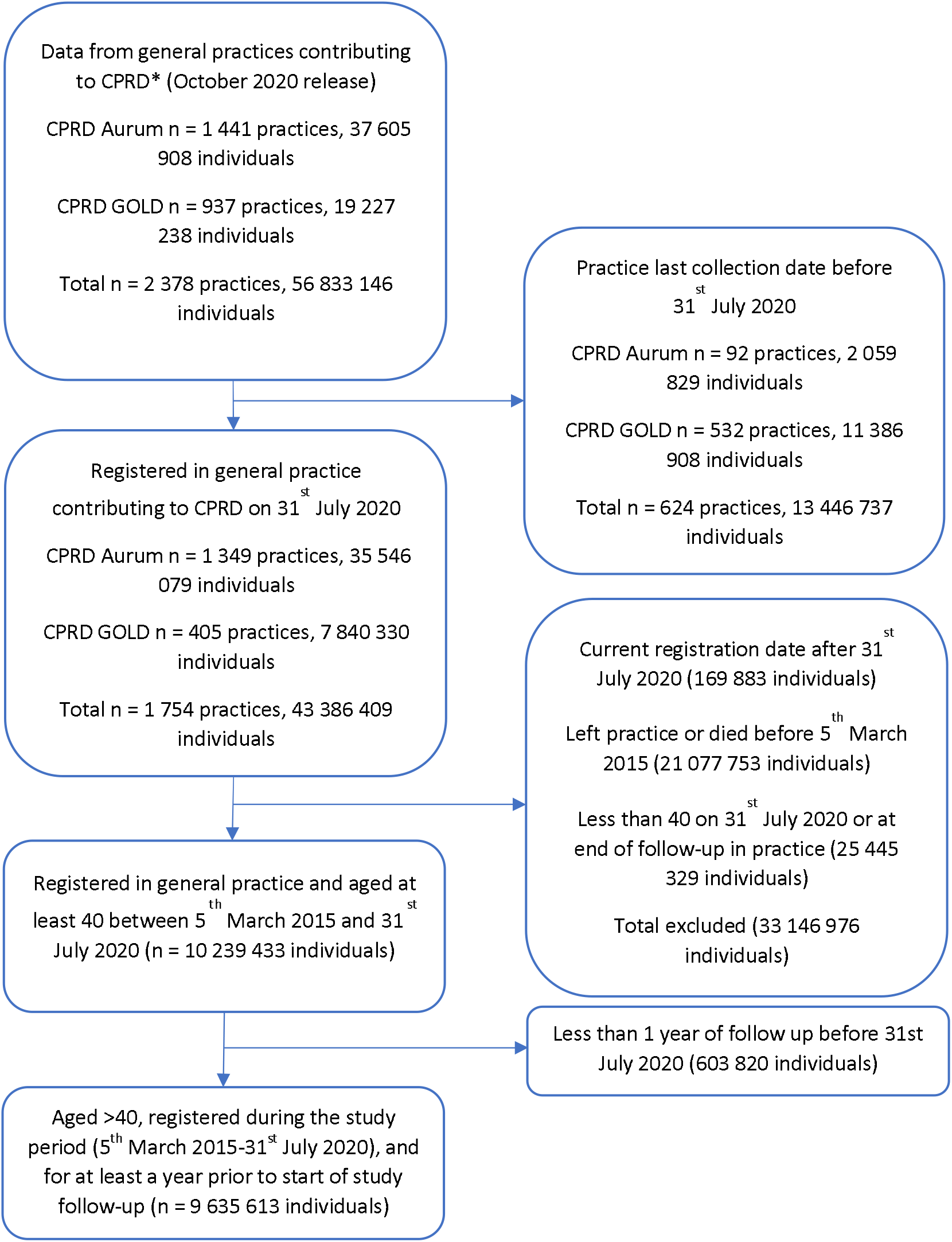
Study cohort selection for primary analysis. Legend: *Practices may contribute to both CPRD GOLD and CPRD Aurum over different time periods. Initial cohort restricted to patients who were permanently registered in the practice with valid registration data (termed “acceptable” by CPRD).

### Overall excess mortality and mortality rate ratio

Figure 2 shows overall patterns of observed, expected and excess mortality for the full study period. We observed 401 deaths per million person-weeks in Wave 1 compared with an estimated 272 (271-272), indicating that 130 (95% CI 129-130) excess deaths occurred per million person-weeks during this period. The relative rate of death in Wave 1 compared to the pre-pandemic period, adjusted for seasonality, year, age, and sex was 1.43 (95% CI 1.40-1.47). Patterns of observed and predicted deaths taking annual and seasonal patterns into account for each comorbidity are described in supplementary appendix p25. Model fit was good for most covariates. Seasonal patterns were consistent, except for people with recent cancer diagnoses where they were less pronounced.

**Figure 2:**
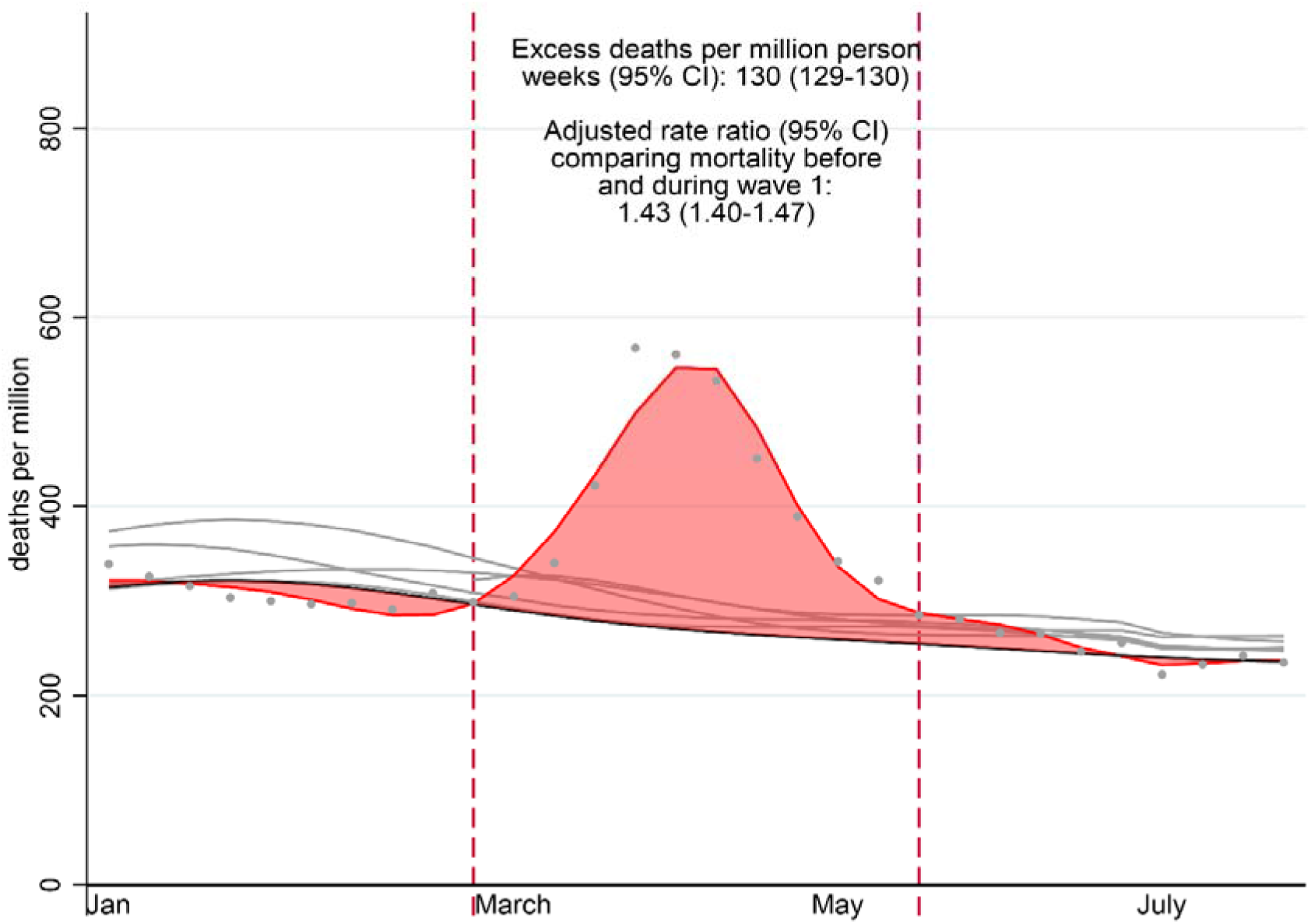
Observed number of deaths from all causes per million person-weeks in 2020 before and during the pandemic and year-specific fitted curves for the years 2015 to 2020. Legend: Solid grey lines – Model fitted numbers of deaths in 2015-2019 (predicted from year and season model restricted to 2015 to 2019) Grey dots – Observed deaths in 2020 Solid red line –Model fitted number of deaths in 2020 using natural cubic spline (with 8 knots per year before the pandemic and an additional 3 knots during the pandemic) Solid black line – Predicted number of deaths in 2020 from year and season model fitted on 2015-2019 data Red shaded area – Excess number of deaths during the pandemic Excess deaths per million patient weeks = Total number of observed deaths in Wave 1 of the pandemic minus predicted number according to the 2015-2019 model Adjusted rate ratio of wave 1 versus pre-pandemic period– Estimated by the model fitted on the full study population adjusted for age, sex, annual and seasonal effects.

### Excess mortality by health and demographic factors

Table 1 describes the population at risk, observed, expected and excess deaths during Wave 1 of the pandemic for each factor. Most deaths occurred among the elderly population and in people who had pre-existing chronic conditions, notably hypertension, chronic kidney disease and dementia.

The number of excess deaths per million-person weeks varied greatly between factors. This is demonstrated by the 100-fold difference between those aged 40-49 and those aged 80 plus years. Numbers of excess deaths per million-person weeks were highest amongst people with dementia (2,693; 95% CI 2,682-2,704), cerebrovascular disease (656; 95% CI 652-660) or cancer diagnosed in the last year (616; 95% CI 603-628), and for people who were underweight (628; 95% CI 620-635).

### Mortality rate ratios by health and demographic factors

Figure 3 displays relative rates of death for each factor before and during Wave 1, adjusted for age, sex, seasonality and year. In most instances, there was strong evidence (p<0.01) of small increases in the association between factors and mortality observed in the pre-pandemic period compared to Wave 1. For example, each five-year increase in age was associated with 1.67 (95% CI 1.67-1.68) times greater rate of death in the pre-pandemic and 1.70 (95% CI 1.69-1.71) times greater rate of death in Wave 1 of the pandemic (p value for interaction between categorical age and the pandemic term <0.01).

**Figure 3:**
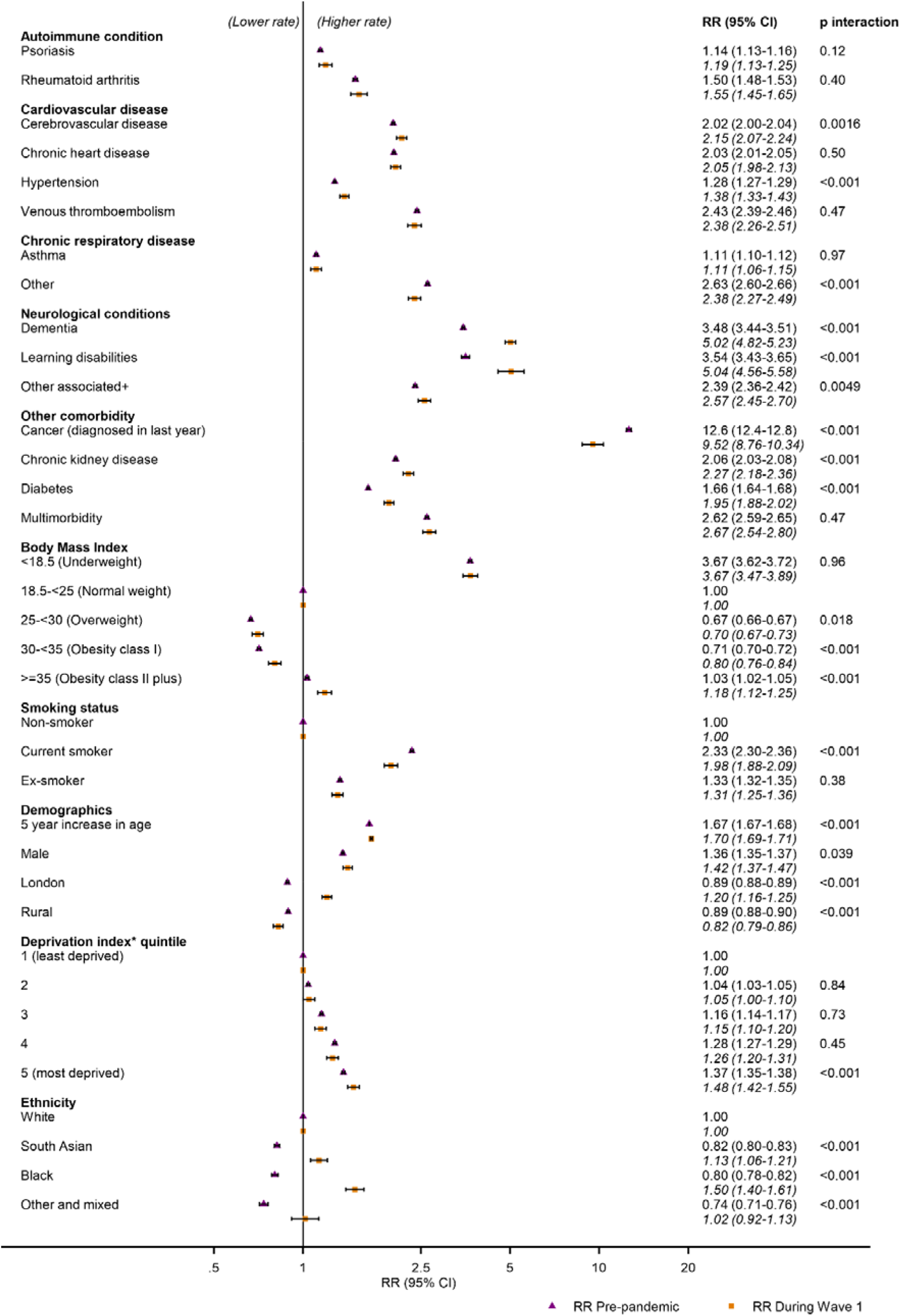
All cause-relative rates of death and 95% confidence intervals by morbidities, health and demographic factors pre-pandemic and during Wave 1 adjusted for age, sex, season and year. Legend: Adjusted for age, sex, season and year. *Carstairs Index. +associated with respiratory viruses. Reference = absence of factor unless otherwise specified. P interaction = P-value for the interaction between the factor/category and Wave 1 of the pandemic

For a minority of factors there were appreciable differences in the relative rate of death between the pre-pandemic and pandemic periods. Pre-pandemic, mortality rates were lower in London compared to other regions of the UK (RR 0.89, 95% CI 0.88-0.89), while in Wave 1 mortality rates were higher in London than in other regions of the UK (1.20, 95% CI 1.16-1.25). Similarly, compared to white ethnic groups, pre-pandemic mortality rates were lower in Black (0.80, 95% CI 0.78-0.82), South Asian (0.81, 95% CI 0.80 - 0.83), and other non-white ethnic groups (0.66, 95% CI 0.64-0.69). However, during Wave 1 of the pandemic, relative rates of death in minority ethnic groups were higher compared to white people (Blacks 1.53, 95% CI 1.43-1.64; South Asians 1.15, 95% CI 1.08-1.23; other ethnicities 1.03, 95% 0.91-1.17).

One of the most striking findings was that there was a substantial increase in the RRs for dementia and learning difficulties in the Wave 1 compared to the pre-pandemic period. Pre-pandemic, people with dementia had a 3.47 (95% CI 3.44-3.51) times higher mortality rate than those without dementia, but this increased to 5.07 (95% CI 4.87-5.28) times higher in Wave 1. The equivalent estimates for learning difficulties were 3.55 (95% CI 3.44-3.51) pre-pandemic, increasing to 4.82 (95% CI 4.35-5.34) during Wave 1.

Cancer diagnosed in the last year was associated with elevated mortality rates both before the pandemic and during Wave 1, but this association was attenuated during Wave 1 (before pandemic: 11.1, 95% CI 10.9-11.3; during Wave 1: 8.37, 95% CI 7.74-9.05).

### Secondary and sensitivity analyses

Supplementary appendix p26 shows relative rates of death for alternative cancer definitions compared with those who never had cancer. RRs were slightly lower during Wave 1 than before the pandemic, except for haematological cancers diagnosed in the last year. Supplementary appendix tables 2 to 5 show RRs by patient or database characteristics. We observed higher RRs before and during Wave 1 of the pandemic for people aged 40-69 compared to people age 70 or older for most risk factors (supplementary appendix p27). When comparing sexes (supplementary appendix p28) and databases (supplementary appendix p29), similar patterns were observed when comparing pre-pandemic RRs to Wave 1 RRs, except for ethnicity and region. Minimal differences were observed when using linked ONS mortality and HES APC data (supplementary appendix p30) except for cancer recently diagnosed, where higher RRs were observed when using the linked data pre-pandemic and multimorbidity where higher RRs were observed both pre-pandemic and during Wave 1.

Supplementary Appendix p31 compares RRs in people of non-white ethnicity versus white ethnicity in London compared to the rest of the UK. The observed increased rates during Wave 1 were attenuated in both London and the rest of the UK compared to the primary analyses which included all UK practices.

Supplementary Appendix p31 shows RRs for missing data categories. RRs for missing categories most closely resembled the baseline (larger) category (white ethnicity, non-smokers and normal weight), therefore indicating that their exclusion may not have led to major biases in the estimated RRs.

## Discussion

### Statement of principal findings

In our very large UK study population, the rate of death by any cause increased by 43% (95% CI 40% to 47%) during Wave 1 of the pandemic compared to that expected based on pre-pandemic levels and trends (2015-19). There were small (+/- 10%) increases in the relative rate of death during wave 1 associated with most of the factors we examined, compared to the pre-pandemic era. For example, chronic heart disease was associated with 2.03 (95% CI 2.01-2.05) times higher rate of death during Wave 1, while pre-pandemic the relative rate was 2.05 (95% CI 1.98-2.13). However, bigger changes in the relative rate of death were seen for people with dementia or learning disabilities, with people in these groups having an approximate 5-fold increased rate of death during Wave 1, compared to people without these conditions, while pre-pandemic this was 3.5 times higher. During Wave 1, we also observed an inversion of the mortality patterns by region and ethnicity: London registered the highest relative rate of death, while pre-pandemic had the lowest; and people of Black or south Asian ethnicity had lower rates of death pre-pandemic, compared to the white population, but markedly increased rates during Wave 1.

### Strengths and weaknesses of the study

A major strength of this study is the use of two very large and well-established datasets of primary care electronic health records. This allowed us to estimate effects for a large and diverse range of chronic conditions, including cardiovascular, respiratory, neurological, and renal diseases, and rarer conditions that would be difficult to study otherwise. CPRD data are of good quality, with high completeness and validity reported for both diagnoses and recorded deaths[24,28]. The availability of data from several years prior to the pandemic permitted us to account for secular and seasonal trends in mortality, and to time-update exposures such as smoking status, obesity, and asthma. Finally, we carried out multiple sensitivity analyses to assess the robustness of the results.

However, this study also has limitations. There may have been misclassification of the vital status of a small number of individuals in some weeks due to imprecise recording of the exact date of death in CPRD. We expect this to have little impact as 98% of the death dates in CPRD are within 30 days of the ONS date of death[24] and our sensitivity analysis using the ONS death date yielded similar results. There is also a potential for misclassification of the exposures, as information may be incomplete (e.g. diagnoses from secondary care not coded in the primary care record) or inaccurate (e.g. patients correctly reporting their smoking behaviour). However, the similar results of the sensitivity analyses including hospital diagnoses in addition to primary care ones suggest minor impact for most conditions. Mortality rate ratios for cancer recently diagnosed and multimorbidity were, however, higher in linked data compared to when primary care data only were used, reflecting known under ascertainment of cancer in primary care data[29]. This was probably exacerbated by the reduced diagnostic activity during Wave 1 reported previously[30]and depicted in our graphs of person weeks contributing over the study period. Thus our main results may underestimate the rates of death in people with cancer because of misclassification, especially during the pandemic. Even though CPRD data are representative of the UK population in terms of age and sex, they include few practices from Eastern England[20]. This may explain differences observed in RRs for ethnicity and region in our sensitivity analyses separating the CPRD GOLD and CPRD Aurum databases. For analyses of health factors, we see no reasons to doubt that the pattern observed elsewhere in the UK would not be applicable. Our models for ethnicity, BMI and smoking included only patients with information on these variables. Our findings from these complete cases analyses are nevertheless valid, under the assumption that missingness for these variables is conditionally independent of the outcome[31].

We should note that the excess mortality in Wave 1 reported in our study cannot be dissociated from the widespread efforts to suppress the virus transmission that involved a national lockdown and imposed social distancing. We cannot assume that the risks observed during Wave 1 would apply to other waves, as population behaviour, risk perception and general health profile of the population most likely have changed.

### Strengths and weaknesses in relation to other studies

There have been few studies of how excess mortality during the Covid-19 pandemic varied by health and wider demographic factors. Recently, de Lusignan et al.[16] reported excess mortality among patients from several demographic and clinical groups in England during Wave 1. Our results are broadly consistent with this previous study, but we are unable to directly compare the magnitude of the excess mortality due to methodological differences: in particular the de Lusignan study used national life table data as an external basis for computing expected deaths in contrast to our use of pre-pandemic mortality within the same population. They were therefore unable to assess whether the relative risk of people with health and demographic factors differed in Wave 1 compared to previous years.

Our finding of an increased relative rate of death during Wave 1 of the pandemic in people with and without dementia and learning difficulties is consistent with cohort studies that have shown that excess mortality was higher in care-home residents in England and Wales, especially those living in care homes catering for older people and those with dementia[32,33], and with ONS data that showed that 30% of all COVID-19-related deaths in England and Wales between March and June 2020 occurred in care homes, and 26% of all COVID-19-related deaths were in people with dementia[34].

Rates of SARS-CoV-2 infection were much higher among those living in care homes than among those living in private homes during Wave 1. This disproportionate exposure to SARS-CoV-2 may explain the increased mortality rate ratio in these two groups of the population. We could not formally test this hypothesis, however, as we did not have data on the type of dwelling (collective vs private), and information on SARS-CoV-2 infection status in Wave 1 was limited by testing capacity.

Going in the other direction from dementia and learning disabilities, the relative rate of death among people with a recent diagnosis of cancer was lower during Wave 1 than it was pre-pandemic. This may be due to a lower risk of SARS-CoV-2 infection in cancer patients, compared to those without cancer consequent upon shielding. On 22 March 2020, the UK Government issued a list of pre-existing clinical conditions that were considered at the time to put people at particularly high risk of serious disease or death if they contracted COVID-19 which included active cancer[35]. In July 2020, the ONS undertook a survey of clinically extremely vulnerable persons in England, concluding that 95% reported either completely or mostly following government shielding guidance[36].

Demographic factors associated with raised risk of exposure to SARS-CoV-2 may also explain the partially interrelated changes in the RR observed during Wave 1 for region and ethnicity. London is a global multicultural city with major international links and likely an important point of entry of imported SARS-CoV-2 infected cases. As the most densely populated region in the UK, with 5,701 people per squared kilometre[37], infection spread fast in London prior to the implementation of control measures[38]. The ethnic inequalities in COVID-19 mortality in Wave 1 in the UK have been reported consistently [7,8,16], and likely related to a complex interaction of factors including a predominance in some public facing occupations (e.g. food retail, healthcare), larger and often multi-generational households, and deprivation. Increases on excess mortality in ethnic minority groups have also been reported in the United States and Sweden[39–41].

For many of the factors we examined, including most morbidities, there was little change in the relative rate of death in Wave 1 compared to pre-pandemic. This is in line with previous research suggesting similar strengths of associations with risk factors for mortality due to COVID-19 and other causes[42], but still striking. This firstly suggests that most of these characteristics are not particularly predictive of exposure to SARS-CoV-2. Beyond this however it can be interpreted as demonstrating that the net effect of COVID-19 in different sub-groups of the population is to simply amplify baseline mortality risk by a constant amount. This is akin to David Spiegelhalter’s observation that the COVID-19 case-fatality age-curve in Wave 1 ran almost perfectly in parallel with the exponential increase with age in the risk of death pre-pandemic [43]. This has been interpreted as showing that COVID-19 has the effect of compressing one’s annual risk of death (whatever that may be) into fewer weeks. This insight, applied to our population study of excess deaths, suggests that the effect of the pandemic has been to accelerate the tempo of underlying mortality rate by a fixed proportional degree. However, the pathophysiological underpinning of this remains unclear and is beyond the scope of this study.

### Meaning of the study and future research

This study has implications for clinical practice, policy, and future research. For most morbidities, the RR did not change very much because of the pandemic, which means that from a clinical perspective pre-pandemic knowledge about the relative frailty associated with different conditions can be reasonably applied in the pandemic situation. However, the high mortality observed in some vulnerable groups, such as those with dementia and learning disabilities (many of whom live in institutions), should be a learning moment for the COVID-19 or other pandemic, and preventive measures should be implemented to avoid the spread of potentially fatal infectious agents. The relative rates of death for characteristics that are strongly affected by population behaviour and the regional epidemiology of the virus may change as the pandemic progresses. Future research may clarify whether there were differences across waves in the UK.

## Data Availability

Individual patient data will not be available. The protocol is available in the supplementary appendix (p2-21) and all code lists are provide at https://doi.org/10.17037/DATA.00002269. All analytical code is available upon request to the corresponding author.

## Ethics statement

This study was approved by the London School of Hygiene & Tropical Medicine Ethics Committee (22680) and the Independent Scientific Advisory Committee for the Medicines and Healthcare products Regulatory Agency database research (20_163R).

## Authors and contributors

DAL and KB conceived the study. HS, BDS, DAL and KB contributed to the study design and the protocol. HS and HC did the data management and have verified the analytical dataset. HS and KB did the statistical analysis. All authors interpreted the findings. HS, HC and DAL wrote the first draft of the manuscript. All authors contributed to subsequent drafts.

HS and HC had full access to all data in the study and accept full responsibility for the work and conduct of the study as guarantors. All authors had access to analysis output and take responsibility for the decision to submit for publication. The corresponding author attests that all listed authors meet the authorship criteria and that no others meeting the criteria have been omitted.

## Transparency statement

The lead authors affirm that the manuscript is an honest, accurate, and transparent account of the study being reported; that no important aspects of the study have been omitted; and that any discrepancies from the study as originally planned have been explained.

## Acknowledgements

KB holds a Wellcome Senior Research Fellowship (220283/Z/20/Z) and also held a Sir Henry Dale fellowship (107731/Z/15/Z, jointly funded by the Wellcome Trust and the Royal Society) during this study. This supported HS, HC and KB’s contribution to this study. DAL’s contribution was partly supported by the Basic Research Program of the National Research University Higher School of Economics. This study is based in part on data from the Clinical Practice Research Datalink obtained under licence from the UK Medicines and Healthcare products Regulatory Agency. The data is provided by patients and collected by the NHS as part of their care and support. The interpretation and conclusions contained in this study are those of the author/s alone. The study was approved by the Independent Scientific Advisory Committee (approval number: 20_163). We thank LSHTM’s electronic health records group, especially Helen McDonald for sharing their code lists with us, and Ketaki Bhate for advice with creating a new psoriasis code list.

## Open Access Statement

This research was funded in whole, or in part, by the Wellcome Trust. For the purpose of open access, the author has applied a CC BY public copyright licence to any Author Accepted Manuscript version arising from this submission.

The Corresponding Author has the right to grant on behalf of all authors and does grant on behalf of all authors, a worldwide licence to the Publishers and its licensees in perpetuity, in all forms, formats and media (whether known now or created in the future), to i) publish, reproduce, distribute, display and store the Contribution, ii) translate the Contribution into other languages, create adaptations, reprints, include within collections and create summaries, extracts and/or, abstracts of the Contribution, iii) create any other derivative work(s) based on the Contribution, iv) to exploit all subsidiary rights in the Contribution, v) the inclusion of electronic links from the Contribution to third party material where-ever it may be located; and, vi) licence any third party to do any or all of the above.

