## Supplementary appendix for "Factors associated with excess all-cause mortality in the first wave of COVID-19 pandemic in the UK: a time-series analysis using the Clinical Practice Research Datalink"

**Protocol**

**Approved protocol**

### INDEPENDENT SCIENTIFIC ADVISORY COMMITTEE (ISAC) PROTOCOL APPLICATION FORM

### PART 1: APPLICATION FORM

***IMPORTANT***

**Both parts of this application must be completed in accordance with the guidance note 'Completion of the ISAC Protocol Application Form', which can be found on the CPRD website (https://cprd.com/research-applications).**

| FOR ISAC USE ONLY | |
|---|---|
| **Protocol No. -** | **Submission date -** |

| GENERAL INFORMATION ABOUT THE PROPOSED RESEARCH STUDY |
|---|
| **1. Study Title (Max. 255 characters including spaces)**<br><br>Excess mortality during the COVID-19 pandemic |
| **2.** Research Area (place '**X**' in all boxes that apply) |

| | | |
|---|---|---|
| Drug Safety | | Economics |
| Drug Utilisation | | Pharmacoeconomics |
| Drug Effectiveness | | Pharmacoepidemiology |
| Disease Epidemiology | X | Methodological |
| Health Services Delivery | | |

**3. Chief Investigator**

| | |
|---|---|
| Title: | Professor |
| Full name: | Krishan Bhaskaran |
| Job title: | Professor |
| Affiliation/organisation: | LSHTM |
| Email address: | |
| CV Number (if applicable): | 156_15CESL |
| Will this person be analysing the data? (Y/N) | Y |

**4. Corresponding Applicant**

| | |
|---|---|
| Title: | Dr |
| Full name: | Helen Strongman |
| Job title: | Research fellow |
| Affiliation/organisation: | LSHTM |
| Email address: | |
| CV Number (if applicable): | 419_15 |
| Will this person be analysing the data? (Y/N) | Y |

**5. List of all investigators/collaborators**

| | |
|---|---|
| Title: | Professor |
| Full name: | Bianca De Stavola |
| Job title: | Professor |
| Affiliation/organisation: | UCL |
| Email address: | |
| CV Number (if applicable): | |
| Will this person be analysing the data? (Y/N) | N |
| Title: | Professor |
| Full name: | David Leon |
| Job title: | Professor |
| Affiliation/organisation: | LSHTM |
| Email address: | |
| CV Number (if applicable): | 614_16S |
| Will this person be analysing the data? (Y/N) | N |
| Title: | Dr |
| Full name: | Christopher Rentsch |
| Job title: | Research fellow |
| Affiliation/organisation: | LSHTM |
| Email address: | |
| CV Number (if applicable): | 176_19 |
| Will this person be analysing the data? (Y/N) | N |
| Title: | Professor |
| Full name: | Ian Douglas |
| Job title: | Professor |
| Affiliation/organisation: | LSHTM |
| Email address: | |
| CV Number (if applicable): | 157_15CESL |
| Will this person be analysing the data? (Y/N) | N |
| Title: | Professor |
| Full name: | Liam Smeeth |
| Job title: | Dean of Faculty |
| Affiliation/organisation: | LSHTM |
| Email address: | |
| CV Number (if applicable): | 045_15CEPSL |
| Will this person be analysing the data? (Y/N) | N |
| Title: | Dr |
| Full name: | Helena Carreira |
| Job title: | Research fellow |
| Affiliation/organisation: | LSHTM |
| Email address: | |
| CV Number (if applicable): | 540_18 |
| Will this person be analysing the data? (Y/N) | Y |

[Add more investigators/collaborators as necessary by copy and pasting a new table for each investigator/collaborator]

**6. Experience/expertise available**

List below the member(s) of the research team who have experience with CPRD data.

| **Name(s):** |
| --- |
| Krishnan Bhaskaran |
| Helen Strongman |
| Christopher Rentsch |
| Ian Douglas |
| Liam Smeeth |

List below the member(s) of the research team who have statistical expertise.

| **Name(s):** |
| --- |
| Krishnan Bhaskaran |
| Bianca De Stavola |

List below the member(s) of the research team who have experience of handling large datasets (greater than 1 million records).

| **Name(s):** |
| --- |
| All |

List below the member(s) of the research team, or supporting the research team, who have experience of practicing in UK primary care.

| **Name(s):** |
| --- |
| Liam Smeeth |

#### ACCESS TO THE DATA

**7. Sponsor of the study**

| | |
|---|---|
| Institution/Organisation: | LSHTM |
| Address: | Keppel Street, London WC1E 7HT |

**8. Funding source for the study**

| | | | |
|---|---|---|---|
| Same as Sponsor? | Yes | Y | No |
| Institution/Organisation: | | | |
| Address: | | | |

**9. Institution conducting the research**

| | | | |
|---|---|---|---|
| Same as Sponsor? | Yes | Yes | No |
| Institution/Organisation: | LSHTM | | |
| Address: | Keppel Street, London WC1E 7HT | | |

**10. Data Access Arrangements**

Indicate with an '**X**' the method that will be used to access the data for this study:

| |
|---|
| Study-specific Dataset Agreement |

| | |
|---|---|
| Institutional Multi-study Licence | X |
| Institution Name | LSHTM |
| Institution Address | Keppel Street, London WC1E 7HT |

Will the dataset be extracted by CPRD?

| | | | |
|---|---|---|---|
| Yes | X | No | X |

If yes, provide the reference number:

We will extract data through the Define and Refine on-line tools. CPRD will extract the BMI, smoking and serum creatinine data from CPRD Aurum based on code lists that we provide (ref CPRD00050632)

**11. Data Processor(s):**

| | |
|---|---|
| Processing | X |
| Accessing | X |
| Storing | X |
| Processing area (UK/EEA/Worldwide) | UK |
| Organisation name | LSHTM |
| Organisation address | Keppel Street, London WC1E 7HT |

| |
|---|
| Processing |
| Accessing |
| Storing |
| Processing area (UK/EEA/Worldwide) |
| Organisation name |
| Organisation address |

[Add more processors as necessary by copy and pasting a new table for each processor]

#### INFORMATION ON DATA

**12. Primary care data** (place '**X**' in all boxes that apply)

| | | | |
|---|---|---|---|
| CPRD GOLD | X | CPRD Aurum | X |

Reference number (if applicable):

**13. Please select any linked data or data products being requested**

**Patient Level Data** (place '**X**' in all boxes that apply)

| | | |
|---|---|---|
| ONS Death Registration Data | X | |
| HES Admitted Patient Care | X | |
| HES Outpatient | | |
| HES Accident and Emergency | | NCRAS Cancer Registration Data |
| HES Diagnostic Imaging Dataset | | NCRAS Cancer Patient Experience Survey (CPES) data |
| HES PROMS (Patient Reported Outcomes Measure) | | NCRAS Systemic Anti-Cancer Treatment (SACT) data |
| CPRD Mother Baby Link | | NCRAS National Radiotherapy Dataset (RTDS) data |
| Pregnancy Register | | NCRAS Quality of Life Cancer Survivors Pilot (QOLP) |
| Mental Health Data Set (MHDS) | | NCRAS Quality of Life Colorectal Cancer Survivors (QOLC) |

**Area Level Data** (place '**X**' in one Practice / Patient level box that may apply)

| **Practice level (UK)** | | **Patient level (England only)** | |
|---|---|---|---|
| Practice Level Index of Multiple Deprivation | | Patient Level Index of Multiple Deprivation | |
| Practice Level Index of Multiple Deprivation (index other than the most recent) | | Patient Level Index of Multiple Deprivation Domains | |
| Practice Level Index of Multiple Deprivation Domains | | Patient Level Carstairs Index for 2011 Census | X |
| Practice Level Carstairs Index for 2011 Census (Excluding Northern Ireland) | X | Patient Level Townsend Score | |
| 2011 Rural-Urban Classification at LSOA level | X | 2011 Rural-Urban Classification at LSOA level | |

Reference / Protocol number (where applicable): 00053616

**14. Are you requesting linkage to a dataset not listed above?**

| | | | |
|---|---|---|---|
| Yes | | No | **X** |

If yes, provide the Non-Standard Linkage reference number:

**15. Does any person named in this application already have access to any of these data in a patient identifiable form, or associated with an identifiable patient index?**

| | | | |
|---|---|---|---|
| Yes | | No | **X** |

If yes, provide further details:

#### VALIDATION/VERIFICATION

**16. Does this protocol describe an observational study using purely CPRD data?**

| | | |
|---|---|---|
| Yes | **X** | No |

**17. Does this protocol involve requesting any additional information from GPs, or contact with patients?**

| | | | |
|---|---|---|---|
| Yes | | No | **X** |

If yes, provide the reference number:

### PART 2: PROTOCOL INFORMATION

**Applicants must complete all sections listed below**

**Applications with sections marked ‘Not applicable’ without justification will be returned as invalid**

| |
|---|
| **A. Study Title (Max. 255 characters, including spaces)**<br><br>Excess mortality during the COVID-19 pandemic |
| **B. Lay Summary (Max. 250 words)**<br><br>Tracking of deaths in the population over time (mortality) is an essential part of monitoring and predicting the likely course of the COVID-19 pandemic. The number of deaths attributed to COVID-19 is problematic as likelihood of stating COVID-19 on the death certificate will have changed over time and may have been inconsistently applied across settings. Excess deaths by week from any cause could provide a more objective and comparable way of assessing the scale of the pandemic and formulating lessons to be learned. It is constructed by comparing the deaths over time since the start of the pandemic to the number expected based on the experience of previous non-pandemic years. Crucially this method has as the outcome all-cause mortality data, sidestepping the problems associated with attribution of cause of death to COVID-19.<br><br>Standalone national mortality data will allow tracking of excess mortality over time, but CPRD uniquely includes timely data on deaths alongside extensive clinical and demographic data, backed up by periodic linkages to national mortality data. This study will exploit this important combination of demographic/clinical and death data to examine how excess mortality during the course of the pandemic differs in individuals with different characteristics (e.g. age, sex, smoking) including pre-existing illnesses (e.g. respiratory disease, heart disease, diabetes). The study will provide key information to inform clinical guidance, resource planning and policies for protecting the most vulnerable. |
| **C. Technical Summary (Max. 300 words)**<br><br>In recognition of limitations of cause-specific death data, estimates of excess mortality are routinely used to estimate and compare deaths due to seasonal influenza epidemics. These are based on national death registration data which allows for stratification by age, sex and geographic region. We aim to replicate these analyses for the COVID-19 pandemic in the UK with further stratification by COVID-19 effect modifiers that can be measured in primary care data.<br><br>Time series methods will be used in our primary analyses of observed mortality during the whole period (pre-COVID-19 and during COVID-19) using generalised linear models with a negative binomial error structure. The main outcome is mortality, measured in primary care data using the CPRD derived death date and the main exposure is the pandemic period (1st March 2020 onwards). The model will initially include exposure, age, sex, and terms to capture seasonality and underlying year-on-year trends. Interaction terms will be used to investigate excess mortality during the pandemic according to demographic, lifestyle-related and comorbidity characteristics. Relative and absolute differences in excess mortality will be described.<br><br>We will compare our primary time series approach to national excess mortality estimates. In a secondary analysis to check conclusions, we will compare weekly mortality with expected mortality based on comparable week-of-year mortality in the years prior to the pandemic (standardised mortality ratio (SMR) approach). We will also explore an approach based on individual-level cohort data, with the pandemic period included as a time-updated variable. In secondary analyses we will use updated linked ONS mortality data when available to repeat the analysis using the ONS death date and HES APC data to improve ascertainment of comorbidities. |

**D. Outcomes to be Measured**

All-cause mortality

**E. Objectives, Specific Aims and Rationale**

*Objective and aims:*

The overall objective is to inform public health policy by providing timely evidence on how key individual-level demographic characteristics and comorbidities affect excess mortality risk over time during the course of the COVID-19 pandemic.

*Specific aims (primary):*

1. Validate approaches/data by estimating all cause excess mortality, by age, sex and co-morbidity grouping, over time by week, during the COVID-19 pandemic in the UK, and compare with similar estimates derived from national death registration data.
2. Use time series methods to investigate how all-cause excess mortality varies by key individual-level characteristics (demographics, lifestyle-related, comorbidities).

*Specific aims (secondary)*

3. Check conclusions from the primary time series analysis using an SMR-based approach that compares observed and expected mortality for given week of the year.

4. Conduct an exploratory cohort analysis using individual-level data to investigate how excess mortality varies by characteristics including specified combinations of comorbidities.

*Rationale:*

Excess deaths by week from any cause could provide the most objective and comparable way of assessing the net impact of the pandemic as it progresses, and avoids the significant problems associated with attributing individual deaths to COVID-19. Use of alternative techniques in secondary and exploratory analyses will allow us to estimate the robustness of our model and potentially extend our analyses to less common comorbidities and multimorbidity groups.

**F. Study Background**

Accurate and consistent estimation of key population parameters is required to inform national and global strategies to mitigate and suppress progression of the coronavirus disease 2019 (COVID-19) pandemic. Essential parameters include mortality due to COVID-19. However, variation in testing and recording of cause of deaths in national death registers between countries, over time and by underlying health conditions makes this challenging[1]. Excess mortality over time may therefore be a more objective and comparable way of assessing the scale of the pandemic and formulating lessons to be learned. It is constructed by comparing the deaths over time in 2020 to the number expected based on the experience of previous non-pandemic years. Crucially this method has as the outcome all-cause mortality, sidestepping the problems associated with attribution of cause of death to COVID-19.

It is possible to estimate excess mortality by age, sex and geographic region using national death registration data although there is a delay in publishing these[1]. These methods have been used to measure excess deaths in previous influenza seasons[2] and are being used for the COVID-19 pandemic[3].

Current methods using death registration data alone do not allow stratified estimation of excess deaths for groups of people with clinical risk factors for COVID-19. CPRD uniquely includes timely data on deaths[4] alongside extensive clinical and demographic data, backed up by periodic linkages to national mortality data. This study will exploit this important combination of demographic/clinical and death data to examine how pre-existing illnesses (e.g. respiratory disease, heart disease, diabetes) and risk factors such as BMI and smoking are associated with excess mortality risk during the course of the pandemic. The study will provide key information to inform clinical guidance, resource planning and policies for protecting the most vulnerable.

**G. Study Type**

Descriptive

**H. Study Design**

For our primary analysis, we will use time series methods modelling weekly counts of deaths through regression models that incorporate seasonal parameters and annual trends and use interaction terms with pandemic indicators to estimate excess mortality after the pandemic and effect modification by patient characteristics. To check the robustness of the results, we will use SMR methods which split the time-series data into pre-COVID-19 and COVID-19 periods and use weekly aggregated data from the former to predict expected mortality rates in the latter.

In exploratory analyses, we will develop individual cohort designs which may allow us to estimate more granular patient characteristics (e.g rare comorbidities and multimorbidity groups). Differences between models will be examined and reported on.

#### I. Feasibility counts

Counts based on April 2020 builds

| | CPRD GOLD | CPRD Aurum |
|---|---|---|
| Alive and registered with up to standard* practice on 1 January 2020 | 3 152 150 | 11 090 586 |
| UK countries represented | All | England and N Ireland |
| English regions represented | All except North East and East Midlands | All |
| Median (IQR) weekly deaths in 2019 | 560 (537-620) | 1657 (1549-1845) |
| January 2019 deaths | 3066 | 8563 |
| January 2020 deaths | 2747 | 8318 |
| February 2020 deaths | 2115 | 6613 |

*Up to standard date only available in CPRD GOLD

#### J. Sample size considerations

We will use all available data in CPRD GOLD and CPRD Aurum to maximise regional coverage and the number and combination of comorbidities that we can study whilst modelling changes on a weekly basis. Data extraction will be minimised by using the CPRD GOLD Define and Refine tools to ascertain covariates of interest only, rather than extracting full clinical data (as discussed with Rebecca Ghosh and Dan Dedman).

Methods for computing sample size and power and not well developed for time series analysis, so we took an empirical approach by combining recent CPRD GOLD denominator data with a simulated pandemic starting in week 9 of the final year which doubles death rates and affects patients with a given comorbidity (simulated to have similar prevalence to cancer) twice as badly. Using the time series approach, the IRR (95% CI) for the pandemic was 1.89 (1.74-2.07) and the comorbidity interaction (1.97 ,1.85-2.08). The confidence intervals included the true values (2.0 and 2.0) with adequate precision. We therefore expect the coverage to be good with this study size, even when estimating interaction terms. 8% of cells had zero denominators in the weekly time series dataset created from these simulated data, necessitating some collapsing of strata, and this would rise further if stratifying by >1 comorbidity, hence the need for maximum data (using both GOLD and Aurum) to enable us to explore joint associations of combinations of covariates, using mutually adjusted models.

We will use the July 2020 build for the initial primary care only analyses because these data are the most timely and updated monthly. Our study period will end on 30 April 2020 as there appears to be a two-month delay in registering deaths in primary care data. Analyses will be updated monthly.

**K. Planned use of linked data (if applicable):**

Additional analyses will be completed when set 19 of the linkage data is released, assuming that ONS linked data coverage will then include the pandemic period. We request access to linked ONS mortality data so that analyses can be repeated using ONS certified death as the outcome and linked HES admitted patient care data to improve ascertainment of ethnicity and comorbidities for stratification in later analyses.

Linked practice level Carstairs quintiles are requested to allow stratification by area-based deprivation in the initial analyses using primary care data only. We also request urban rural data as COVID-19 infections and mortality will be affected by deprivation, population density and distance from hospital. Patient level Carstairs quintiles are requested for the linked analysis.

Risk of identification of practice or patient location will be mitigated by restricting users who have access to the raw CPRD and linked data to Helen Strongman and Krishnan Bhaskaran who understand the increased risk of identification with these data and have completed mandatory data protection training at LSHTM. This will be implemented by password protecting folders where data are stored in accordance with LSHTM IT policy. Patient and practice identifiers will be pseudonymised in datasets released to other investigators.

Analyses using linked data will be restricted to English practices and patients who are eligible for linkage, and to the time periods of linked data coverage.

This study will use demographic/clinical and death data available in CPRD primary care and linked data sources to examine how pre-existing illnesses (e.g. respiratory disease, heart disease, diabetes) and risk factors such as BMI and smoking are associated with excess mortality risk during the course of the COVID-19 pandemic in the UK. The findings will provide key information to inform clinical guidance, resource planning and policies for protecting the most vulnerable people in the UK, including in England and Wales, during the pandemic.

**L. Definition of the Study population**

Data from practices that have contributed to both CPRD GOLD and CPRD Aurum will be removed from the CPRD Aurum cohort to maximise the power of analyses that can only be completed in CPRD GOLD (i.e. stratification by number of people in household).

1) Time series and SMR approaches

The study population will be the adult population of all practices with continuous follow-up from 1st March 2014 and with last data collection dates no more than 2 months before the date of the most recent CPRD build (“time-series cohort”).

2) Cohort approach

The population under investigation will be the adult population contributing CPRD follow-up at any time after 1st March 2014.

**M. Selection of comparison group(s) or controls**

For the time series and SMR approach, the comparison is between observed aggregated mortality before and during the pandemic, investigated using respectively interaction terms with pandemic indicators, or comparisons with expected mortality modelled using pre-COVID-19 weekly aggregated data.

For the cohort approach, the comparison is between mortality during the pandemic period and mortality during the pre-pandemic period where information is kept at the individual-level (as opposed to aggregated).

**N. Exposures, Outcomes and Covariates**

The main exposure of interest is the Covid-19 pandemic starting from 1st March 2020.

The main outcome is mortality, identified from primary care data using the CPRD derived death date. In secondary analyses we will use updated linked ONS mortality data when available to repeat the analysis using the ONS death date.

Covariates of interest for stratification are:

- Geographical region
- Age (initial time series counts to be generated in 5 year categories, then collapsed further for analysis)
- Sex
- Smoking status (current, never, ex)
- BMI group (WHO groupings)
- Ethnicity (White, South Asian, Black, Mixed Other)
- Number of individuals in a household(CPRD GOLD only)
- Care home residency (CPRD GOLD only)
- History of comorbidities including those relevant to government "vulnerable groups" policy*:
    - Asthma
    - Chronic respiratory disease (other than asthma)
    - Cardiovascular disease
    - Diabetes
    - Cancer
    - Chronic liver disease
    - Chronic neurological diseases
    - Chronic kidney disease
    - Organ transplant recipients
    - Problems with spleen
    - On immunosuppressive drugs
    - Hypertension
    - Autoimmune disease
    - Endocrine disease (including thyroid others)
    - Multimorbidity

We will use code lists and definitions developed by the LSHTM Health Protection Research Unit on Immunisation where available, and where needed develop new code lists using systematic dictionary keyword searches, manual review, and clinical sign-off; these will be published online and linked in research outputs.

*Current UK government guidance on who should be considered vulnerable has been largely based on policies developed for previous epidemic respiratory viruses, notably influenza. Understanding of Covid-19 risk factors is improving rapidly. We will submit a protocol amendment should new Covid-19 risk factors that can be measured using the available data emerge during the study.

#### O. Data/ Statistical Analysis

To ensure our conclusions are robust, several approaches will be taken (with different strengths and limitations). We would expect the overall patterns and conclusions of the analyses to be similar. Any discrepancies in the patterns or results will be investigated and reported. In each case we will use data up to two months before the CPRD build date (i.e. data to end April 2020, from the build released in early July 2020) and analyses will be updated monthly.

##### 1) Primary analysis - time series approach

We will derive weekly counts of numbers of deaths in contributing practices (see section L "Study Population") from the start of the study period to the end of the study period. Population denominator sizes (number under active follow-up) will also be extracted. We will extract these counts overall, and stratified by age, sex, region and other factors (see section N "Covariates"). Where these factors are time-varying, counts will reflect changes over time.

We will fit generalised linear models with a negative binomial error structure to model the counts of deaths, taking account of variable denominator sizes [approach we took in Matthews et al media and statins paper to deal with variable denominators, BMJ2016[5]]. The model will initially include the outcome (mortality), exposure (2020 pandemic period vs pre-pandemic), age, sex, interactions between the exposure and age and sex, Fourier terms (a harmonic function of calendar week) to capture seasonality, a continuous function of calendar year (linear, or higher order polynomial if evidence of non-linearity) to capture any underlying trends, and if relevant dummy indicators to capture extreme flu seasons. When investigating excess mortality within subgroups, further exposure covariate stratification factors will be added individually, and in combinations where there is sufficient information. Interactions between the exposure and stratification factors will provide estimates of the relative effect of the pandemic in strata.

##### 2) Secondary analysis - SMR-based approach

We will use the stratified weekly counts generated for the time series approach, and collapse years into two periods (all pre-pandemic vs 2020 pandemic). Age/sex/comorbidity-specific expected weekly death rates for the pandemic period will be generated using the pre-pandemic data, and compared with observed weekly deaths in the pandemic period. Risk factors will be analysed one at a time, and combined where numbers allow. Standardised mortality ratios will be calculated as observed / expected and 95% confidence intervals will be generated from a saturated Poisson model.

##### 3) Secondary analysis - Cohort approach

Using data from March 1$^{st}$ 2014 to the end of the study period, we will fit a proportional hazards model for time to death on an underlying age timescale (with extensions if proportionality of the hazards assumption is not met). Individuals will enter the analysis 1 year after entry into CPRD to allow classification of comorbidities. Time-updated calendar month and year will be included in the model, alongside a time-updated indicator for the pandemic period, and individual-level covariates. Interactions between the pandemic indicator and covariates will be added one at a time initially and in combinations, where possible, to describe differences in excess mortality between risk factor groups.

<u>Validation of approaches / data</u>

We will compare estimates of all cause excess age/sex-specific mortality over time by week during the course of the COVID-19 pandemic with similar estimates derived from national death registration data.

**P. Plan for addressing confounding**

This is not a hypothesis testing/causal modelling study, so confounding per se is not an issue. The role of covariates will be explored by stratification in the main analyses and by adjustment and stratification in the exploratory individual cohort analyses.

**Q. Plans for addressing missing data**

Analyses stratified by BMI, smoking and ethnicity are likely to have missing data. In our primary analyses, we will create binary yes/no variables for obesity, ever smoked, white ethnicity assuming that individuals with missing data are not obese, have never smoked and are white. We will conduct sensitivity analyses restricted to the study population with data available, with appropriate assessment of possible selection bias affecting the results. We will also conduct sensitivity analyses whereby missing obesity, smoking and ethnicity data are randomly recoded as taking any value other than the reference to assess the impact of misclassification bias.

**R. Patient or user group involvement**

There is no patient or user group involvement. This is a broad study and does not focus on a specific disease group, and the requirement to proceed on short timescales to inform government policy precludes consultation with user groups.

**S. Plans for disseminating and communicating study results, including the presence or absence of any restrictions on the extent and timing of publication**

Results will be released at the earliest opportunity in order to inform policy in a timely way, including release of data on a preprint server. Peer review and formal publication will proceed in due course

**Conflict of interest statement:**

There are no conflicts of interest to declare.

#### T. Limitations of the study design, data sources, and analytic methods

**Data sources**

Our initial analyses will use primary care data only. This is the mostly timely data available to us as it is updated monthly and we believe that deaths are recorded within two months. Small lags have been reported between the date of death in primary care records compared to death registration data. We will therefore validate our approach through comparison with nationally reported excess mortality data.

Measurement of covariates such as ethnicity and conditions treated in hospital will also be affected. We will acknowledge these limitations and repeat our analyses using linked HES APC and mortality data when available.

**Analytic methods**

Time series and SMR analyses cannot confirm a causal link between the pandemic and the observed changes in mortality, or disentangle direct impacts of Covid-19 from indirect impacts of the pandemic and control measures. The design accounts for individual level factors such as smoking and obesity that remain fairly constant or change relatively slowly overtime within the population, but it is possible that other external factors played a role in the observed changes (e.g. influenza, air pollution). In the first few months of the pandemic, the impact of these changes are likely to be minimal compared to the impact of COVID-19.

Through stratification within our time series and SMR analyses, we will be able to show how the effect size differs between people with and without individual risk factors and two-way combinations of comorbidities. We will not be able to fully explore the impact of multimorbidity. However, through exploration of individual cohort methods, we hope to be able to estimate independent effects of risk factors, assuming no residual confounding.

**List of Appendices**

**Amendment – 21 October 2020**

Explanation of change: We have added Helena Carreira as a collaborator for this study. Helena will have access to the data which requires the following change to section J.

Before: Risk of identification of practice or patient location will be mitigated by restricting users who have access to the raw CPRD and linked data to Helen Strongman and Krishnan Bhaskaran who understand the increased risk of identification with these data and have completed mandatory data protection training at LSHTM. This will be implemented by password protecting folders where data are stored in accordance with LSHTM IT policy. Patient and practice identifiers will be pseudonymised in datasets released to other investigators.

After: Risk of identification of practice or patient location will be mitigated by restricting users who have access to the raw CPRD and linked data to Helen Strongman, Krishnan Bhaskaran and Helena Carreira who understand the increased risk of identification with these data and have completed mandatory data protection training at LSHTM. This will be implemented by password protecting folders where data are stored in accordance with LSHTM IT policy. Patient and practice identifiers will be pseudonymised in datasets released to other investigators.

**Changes to approved protocol**

This manuscript reports findings from the primary time series analysis. We compared this to the SMR-based approach early in the study and found that using both approaches did not add to the study. We have not attempted the proposed exploratory cohort analysis using individual-level data to investigate how excess mortality varies by characteristics including specified combinations of comorbidities.

To avoid problems with regression model convergence due to low numbers of deaths, we changed the study population to include people aged >= 40 and have not reported findings for chronic liver disease, systemic lupus erythematosus, or immunosuppression.

We started individual follow-up from 1 year after registration in the practice to avoid missing information about chronic morbidities and health status that may be added after the patient registered in the practice.

#### Risk factor definitions

| Risk factor | Definition | Derivation |
|---|---|---|
| **Demographic** | | |
| Age | 5-year age groups from 40-<45 to ≥ 980 | 01/07/birthyear (Exact date of birth not collected by CPRD to maintain the de-identified nature of the data) |
| Sex | Male, Female | Recorded in primary care record |
| Region | Value to indicate where in the UK the practice is based. The region denotes the former Strategic Health Authority for practices within England, and the country i.e. Wales, Scotland, or Northern Ireland for the rest | Provided by CPRD |
| Practice area-based deprivation | Carstairs Index quintile used as proxy for socio-economic status (1 least deprived, 2, 3, 4, 5 most deprived) | Carstairs Index quintile identified by CPRD through linkage to practice postcode |
| Patient area-based deprivation | Replaces practice area-based deprivation in sensitivity analysis using linked data | Carstairs Index quintile identified by CPRD through third party linkage to patient postcode |
| Rural-Urban | Rural or Urban | Identified by CPRD through linkage of Rural Urban Classifications for England & Wales, Scotland and Northern Ireland to practice postcode. Mixed Urban-Rural in Northern Ireland reclassified as Urban. |
| Ethnicity | 5 categories, as defined in the Census: White; Mixed or multiple ethnic groups; Asian or Asian British; Black, African, Caribbean or Black British; Other. | Derived from information in the patient clinical records in analyses of primary care data. In analyses of primary care data linked to hospital data, ethnicity variable was further supplemented with the information in the hospital record.<br><br>(https://academic.oup.com/jpubhealth/article/36/4/684/1529704) |
| **Morbidity** | | |
| **Cardiovascular** | | |
| Chronic heart disease | Includes ischaemic heart disease, congenital heart disease, heart failure/cardiomyopathy, valve diseases, and primary hypertension | Any coded previous clinical diagnosis, major intervention for, or clinical review for chronic heart disease. Codes from primary and secondary care included in the sensitivity analysis. |
| Cerebrovascular disease | Stroke or transient ischaemic attack | Any coded previous clinical diagnosis, major intervention for, or clinical review for cerebrovascular disease. Codes from primary and secondary care included in the sensitivity analysis. |
| Venous thrombo-embolism | Venous thrombo-embolism | Any coded previous clinical diagnosis, major intervention for, or clinical review for venous thrombo-embolism. Codes from primary and secondary care included in the sensitivity analysis. |
| Hypertension | Hypertension | Diagnosis code or two abnormal blood pressure records (systolic blood pressure ≥140 mmHg or diastolic blood pressure ≥90 mmHg) within a 2-year period. Assumed to be chronic. (https://www.ncbi.nlm.nih.gov/pmc/articles/PMC5072168/) |
| **Respiratory** | | |
| Asthma | Current asthma diagnosis | Specific record of asthma diagnosis in last 3 years. (Validation of asthma recording in the Clinical Practice Research Datalink (CPRD) \| BMJ Open). Codes from primary and secondary care included in the sensitivity analysis. |
| Other chronic respiratory condition | Chronic obstructive pulmonary disease (COPD), emphysema, bronchitis, | Any previous coded diagnosis or clinical review. Codes from primary and secondary care included in the sensitivity analysis. |

| | | |
|---|---|---|
| | cystic fibrosis, or fibrosing interstitial lung diseases. | |
| **Autoimmune** | | |
| Rheumatoid Arthritis | Rheumatoid arthritis | Any previous coded diagnosis or clinical review |
| Lupus erythematosus | Lupus erythematosus | Any previous coded diagnosis or clinical review. Excluded from analysis due to problems with model convergence. |
| Psoriasis | Psoriasis | Any previous coded diagnosis or clinical review |
| **Neurological** | | |
| Learning / intellectual disability | Learning / intellectual disability | Any previous coded diagnosis or clinical review. Codes from primary and secondary care included in the sensitivity analysis. |
| Dementia | Dementia | Any previous coded diagnosis or clinical review. Codes from primary and secondary care included in the sensitivity analysis. |
| Other chronic neurological condition associated with respiratory infection | Parkinson's disease, multiple sclerosis, motor neurone disease, cerebral palsy, quadriplegia, progressive cerebellar disease, Huntington's, poliomyelitis, and myasthenia | Any previous coded diagnosis or clinical review. Codes from primary and secondary care included in the sensitivity analysis. |
| **Other** | | |
| Chronic liver disease | Cirrhosis, oesophageal varices, biliary atresia and chronic hepatitis. | Any previous coded diagnosis of or clinical review. Excluded from analysis due to problems with model convergence. |
| Chronic Kidney Disease (CKD) | CKD, history of dialysis or renal transplant. Or with estimated glomerular filtration rate (eGFR) to classify CKD stage. | Any previous coded diagnosis of or clinical review of CKD, dialysis, or renal transplant. Any previous serum creatinine record that allows the calculation of an estimated glomerular filtration rate (eGFR) to classify CKD stage 3a-5.<br><br>Codes from primary and secondary care included in the sensitivity analysis. |
| Diabetes | Diabetes mellitus excluding gestational diabetes | Any previous coded diagnosis of or clinical review. Codes from primary and secondary care included in the sensitivity analysis. |
| Recently diagnosed cancer | All first malignant tumours diagnosed within the last year | First ever malignant cancer diagnosis diagnosed in the last year (time-updating). Not counted if there is a previous record of a historical or secondary malignancy. Codes from primary and secondary care included in the sensitivity analysis. |
| Permanent immunosuppression | HIV, solid organ transplant or other permanent immunosuppression (such as genetic conditions compromising immune function). | Any previous coded diagnosis of or clinical review. Included in Hospital Episode Statistics (HES) sensitivity analysis. Excluded from analysis due to problems with model convergence. |
| **Multimorbidity** | | |
| Multimorbidity | More than one condition of the following: asthma, other chronic respiratory disease, chronic heart disease, chronic kidney disease, chronic liver disease, chronic neurological disease, diabetes or permanent immunosuppression | Counted from diagnosis of second condition within list. |
| **Health Indicators** | | |
| Smoking Status | Non-smoker, smoker, ex-smoker | Previous structured or coded record of smoking status (time updating). Non-smokers recategorised as ex-smoker if there was a previous record of smoking |
| Body Mass Index (BMI) | Categorised based on WHO categories for Caucasian populations | Derived from weight and height measures (BMI category time updating). (https://www.ncbi.nlm.nih.gov/pubmed/24038008) |

| Cancer (secondary analyses) | | |
|---|---|---|
| Haematological Cancer | Lymphoma, multiple myeloma and leukaemia, and other haematological cancers | First ever coded record for malignant cancer identifies haematological malignancy. Not counted if there is a previous record of a historical or secondary malignancy. Separate variables for:<br>- Above record recorded in last year (time-updating)<br>- Above record recorded 1 to 5 years ago (time-updating)<br>- Above record recorded more than 5 years ago |
| Non-haematological Cancer | All other malignant cancers | First ever coded record for malignant cancer identifies non-haematological malignancy. Not counted if there is a previous record of a historical or secondary malignancy. Separate variables for:<br>- Above record recorded in last year (time-updating)<br>- Above record recorded 1 to 5 years ago (time-updating)<br>- Above record recorded more than 5 years ago |

#### Model Checking

##### (A) Millions of person-weeks contributed over study period

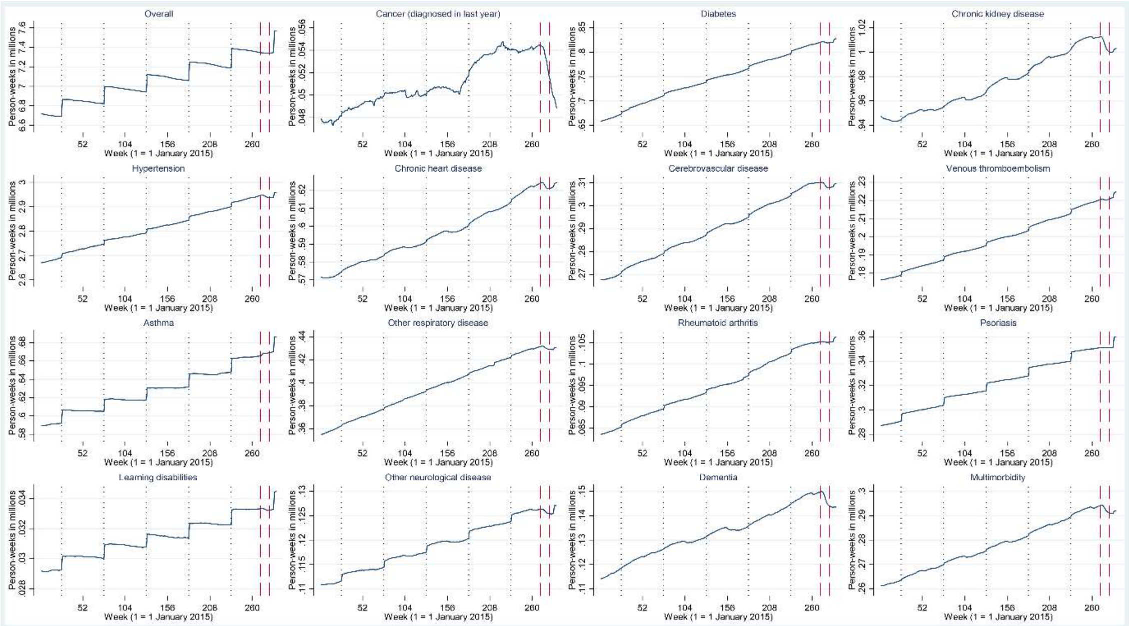

**(B) Observed and predicted deaths per million person-weeks for full study period**

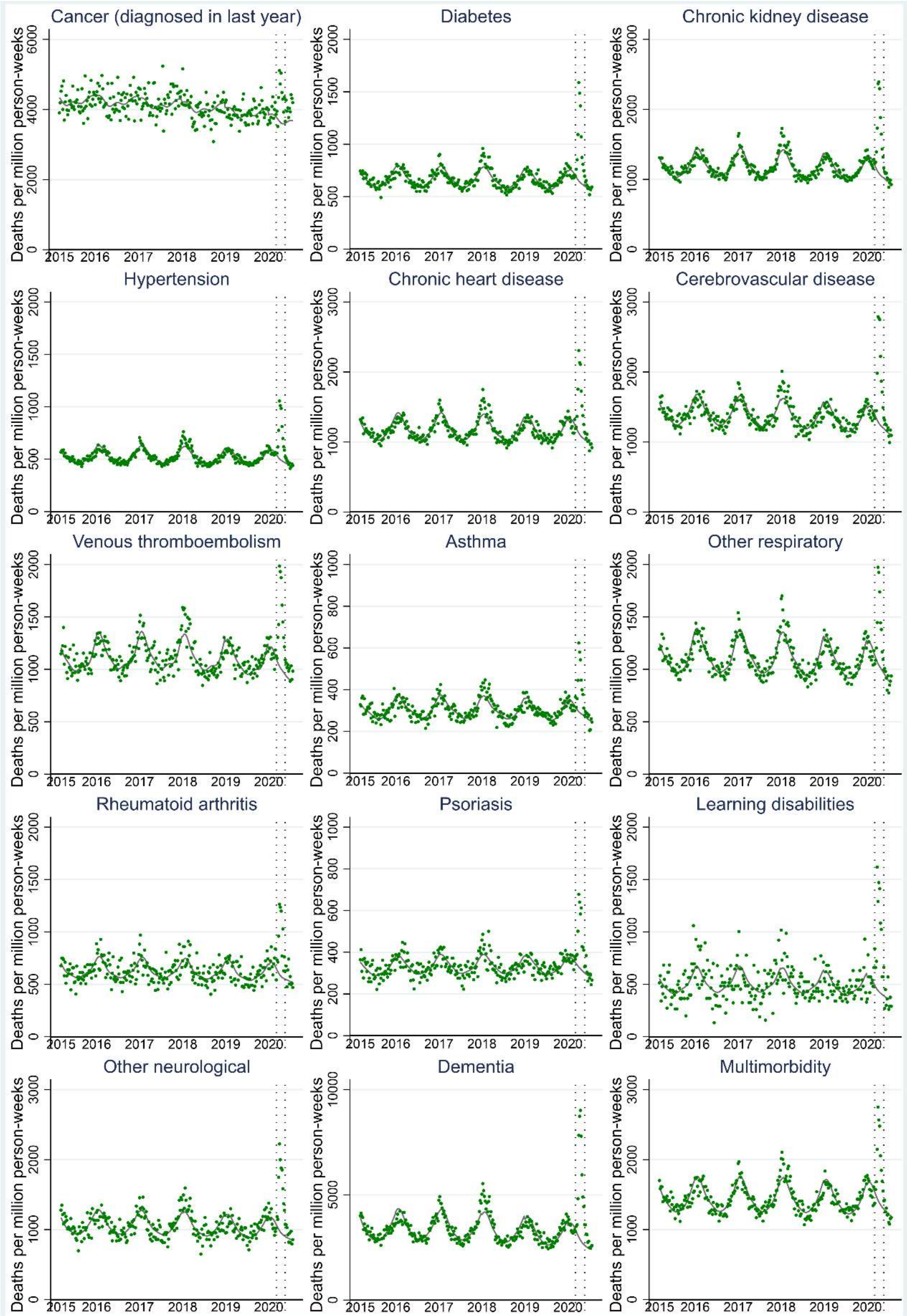

Legend: green dots = observed deaths per million, solid grey line= predicted deaths from year season model restricted to pre-pandemic period, dotted grey lines = start and end of wave 1 (5th March to 27th May 2020). Y axis scale differs for each graph.

#### Secondary analyses

##### Mortality rate ratios comparing all cause death in people with cancer compared to people without cancer before the pandemic and during Wave 1

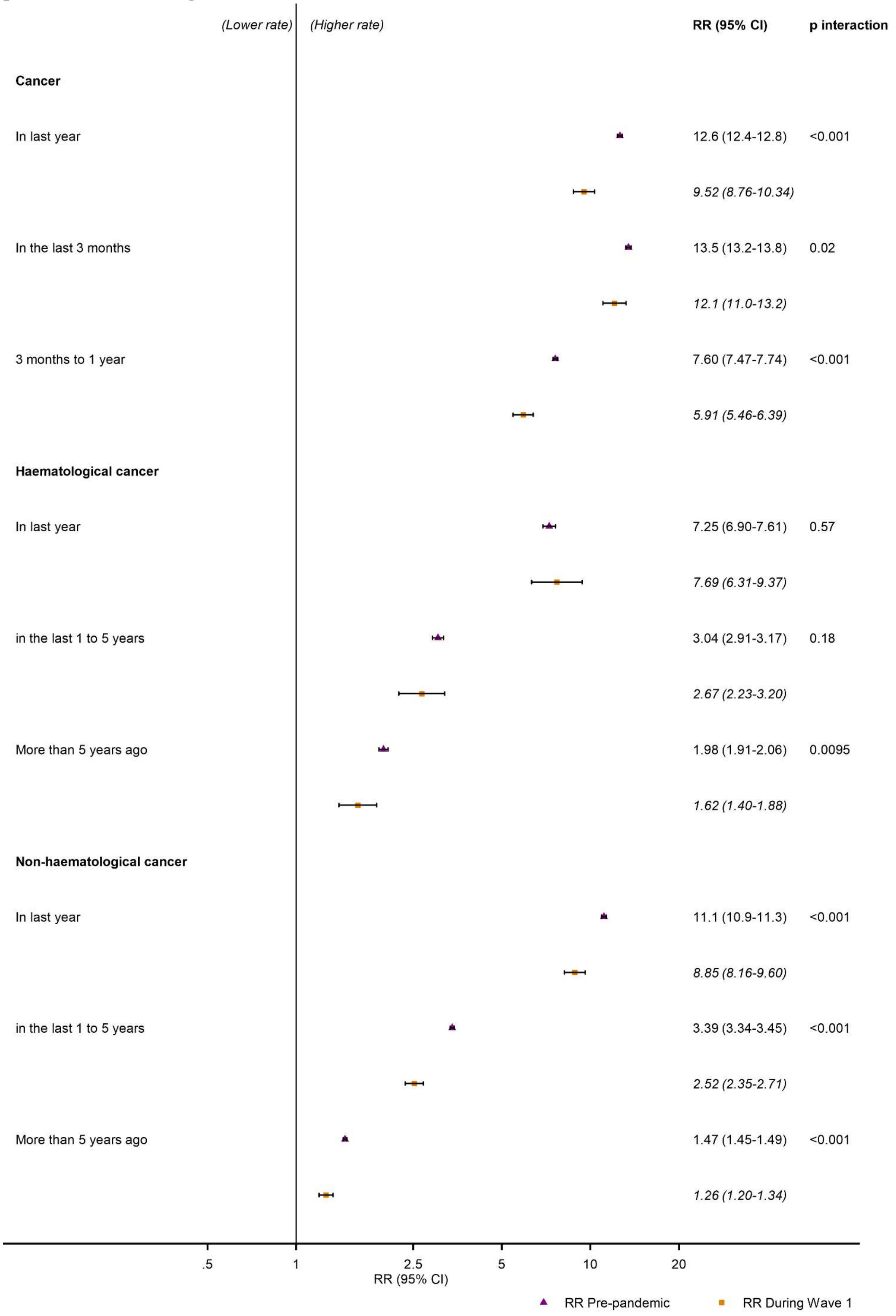

Legend: Adjusted for age, sex, season and year. The result for cancer in the last year replicates the main analysis.

**Sensitivity analyses: All-cause relative rates of death and 95% confidence intervals by morbidities, health and demographic factors pre-pandemic and during Wave 1 adjusted for age, sex, season and year**

**(A) By Age group**

| | Pre-pandemic | | During Wave 1 | |
|---|---|---|---|---|
| | **40 to 69** | **70 or older** | **40 to 69** | **70 or older** |
| **DEMOGRAPHICS** | | | | |
| **Age** | | | | |
| 5-year increase in age | 1.57 (1.57-1.58) | 1.82 (1.81-1.83) | 1.58 (1.55-1.61) | 1.88 (1.85-1.92) |
| **Sex** | | | | |
| Female | 1.00 | 1.00 | 1.00 | 1.00 |
| Male | 1.45 (1.43-1.47) | 1.32 (1.31-1.33) | 1.56 (1.48-1.64) | 1.36 (1.30-1.42) |
| **Carstairs deprivation index quintile** | | | | |
| 1 (least deprived) | 1.00 | 1.00 | 1.00 | 1.00 |
| 2 | 1.07 (1.04-1.09) | 1.04 (1.02-1.05) | 1.12 (1.02-1.23) | 1.03 (0.98-1.09) |
| 3 | 1.26 (1.23-1.29) | 1.13 (1.11-1.14) | 1.22 (1.11-1.33) | 1.13 (1.08-1.19) |
| 4 | 1.50 (1.46-1.53) | 1.21 (1.20-1.23) | 1.46 (1.33-1.59) | 1.21 (1.15-1.27) |
| 5 (most deprived) | 1.63 (1.60-1.67) | 1.28 (1.27-1.30) | 1.80 (1.65-1.96) | 1.39 (1.32-1.46) |
| **Ethnicity** | | | | |
| Black | 0.82 (0.79-0.86) | 0.79 (0.77-0.82) | 1.59 (1.42-1.78) | 1.45 (1.33-1.58) |
| Other and mixed | 0.65 (0.61-0.68) | 0.80 (0.77-0.83) | 0.93 (0.78-1.11) | 1.08 (0.95-1.24) |
| South Asian | 0.71 (0.68-0.73) | 0.88 (0.86-0.90) | 1.10 (0.99-1.23) | 1.15 (1.06-1.25) |
| White | 1.00 | 1.00 | 1.00 | 1.00 |
| **Region** | | | | |
| London | 0.85 (0.84-0.87) | 0.90 (0.89-0.91) | 1.18 (1.11-1.26) | 1.21 (1.15-1.26) |
| Other | 1.00 | 1.00 | 1.00 | 1.00 |
| **Urban Rural** | | | | |
| Rural | 0.79 (0.78-0.81) | 0.92 (0.92-0.93) | 0.71 (0.66-0.77) | 0.85 (0.81-0.89) |
| Urban | 1.00 | 1.00 | 1.00 | 1.00 |
| **HEALTH BEHAVIOURS / INDICATORS** | | | | |
| **Body Mass Index** | | | | |
| <18.5 (Underweight) | 5.89 (5.74-6.04) | 3.19 (3.14-3.24) | 5.59 (5.01-6.23) | 3.15 (2.96-3.35) |
| 18.5-<25 (Normal weight) | 1.00 | 1.00 | 1.00 | 1.00 |
| 25-<30 (Overweight) | 0.65 (0.64-0.66) | 0.67 (0.66-0.68) | 0.76 (0.71-0.82) | 0.67 (0.64-0.71) |
| 30-<35 (Obesity class I) | 0.72 (0.70-0.73) | 0.70 (0.69-0.71) | 0.93 (0.86-1.01) | 0.74 (0.70-0.79) |
| >=35 (Obesity class II plus) | 1.15 (1.13-1.18) | 0.95 (0.94-0.97) | 1.64 (1.52-1.77) | 0.96 (0.89-1.02) |
| **Smoking status** | | | | |
| Current smoker | 3.40 (3.34-3.46) | 1.85 (1.82-1.88) | 2.90 (2.69-3.12) | 1.56 (1.46-1.66) |
| Ex-smoker | 1.67 (1.65-1.70) | 1.21 (1.20-1.22) | 1.63 (1.53-1.74) | 1.19 (1.13-1.25) |
| Non-smoker | 1.00 | 1.00 | 1.00 | 1.00 |
| **MORBIDITY** | | | | |
| **Autoimmune condition** | | | | |
| Lupus erythematosus | 2.19 (2.01-2.38) | 1.47 (1.39-1.56) | 1.33 (0.87-2.05) | 1.17 (0.92-1.48) |
| Psoriasis | 1.26 (1.23-1.30) | 1.10 (1.09-1.12) | 1.28 (1.16-1.41) | 1.15 (1.08-1.22) |
| Rheumatoid arthritis | 1.70 (1.63-1.77) | 1.46 (1.44-1.49) | 2.01 (1.74-2.32) | 1.43 (1.33-1.54) |
| **Cardiovascular disease** | | | | |
| Cerebrovascular disease | 3.17 (3.10-3.24) | 1.84 (1.82-1.86) | 3.24 (2.99-3.52) | 1.91 (1.83-2.00) |
| Chronic heart disease | 2.98 (2.93-3.03) | 1.80 (1.78-1.81) | 3.06 (2.86-3.27) | 1.75 (1.68-1.82) |
| Hypertension | 1.57 (1.55-1.60) | 1.14 (1.13-1.15) | 1.82 (1.73-1.92) | 1.16 (1.11-1.21) |
| Venous thromboembolism | 4.99 (4.89-5.09) | 1.80 (1.78-1.83) | 4.64 (4.29-5.01) | 1.74 (1.65-1.84) |
| **Chronic respiratory disease** | | | | |
| Asthma | 1.28 (1.25-1.30) | 1.05 (1.04-1.06) | 1.28 (1.19-1.38) | 1.03 (0.98-1.09) |
| Other | 4.15 (4.08-4.22) | 2.16 (2.13-2.18) | 3.72 (3.49-3.98) | 1.89 (1.79-1.99) |
| **Neurological conditions** | | | | |
| Dementia | 6.85 (6.64-7.06) | 3.25 (3.22-3.29) | 7.77 (7.01-8.62) | 4.73 (4.51-4.96) |
| Learning disabilities | 4.54 (4.36-4.72) | 2.60 (2.48-2.74) | 6.46 (5.66-7.37) | 3.90 (3.33-4.56) |
| Other associated with respiratory infections | 3.69 (3.60-3.78) | 2.15 (2.12-2.18) | 3.84 (3.48-4.25) | 2.29 (2.17-2.42) |
| **Other comorbidity** | | | | |
| Cancer (diagnosed in last year) | 26.4 (26.0-26.9) | 6.85 (6.73-6.97) | 20.3 (18.7-21.9) | 5.01 (4.63-5.43) |
| Chronic kidney disease | 3.42 (3.36-3.47) | 1.70 (1.68-1.71) | 3.97 (3.72-4.23) | 1.76 (1.69-1.83) |
| Diabetes | 2.27 (2.23-2.30) | 1.46 (1.45-1.48) | 2.89 (2.73-3.07) | 1.61 (1.54-1.68) |
| Multimorbidity | 4.77 (4.68-4.86) | 2.12 (2.09-2.14) | 5.18 (4.82-5.57) | 1.99 (1.89-2.10) |

**(B) By Sex**

| | Pre-pandemic | | During Wave 1 | |
|---|---|---|---|---|
| | **Male** | **Female** | **Male** | **Female** |
| **DEMOGRAPHICS** | | | | |
| **Age** | | | | |
| 5-year increase in age | 1.00 | 1.00 | 1.01 (1.01-1.02) | 1.03 (1.02-1.03) |
| **Carstairs deprivation index quintile** | | | | |
| 1 (least deprived) | 1.00 | 1.00 | 1.00 | 1.00 |
| 2 | 1.05 (1.03-1.06) | 1.04 (1.02-1.05) | 1.08 (1.01-1.15) | 1.02 (0.95-1.08) |
| 3 | 1.16 (1.14-1.18) | 1.15 (1.13-1.17) | 1.18 (1.11-1.25) | 1.11 (1.05-1.19) |
| 4 | 1.30 (1.28-1.32) | 1.26 (1.24-1.28) | 1.31 (1.23-1.39) | 1.20 (1.13-1.28) |
| 5 (most deprived) | 1.39 (1.37-1.41) | 1.34 (1.32-1.36) | 1.54 (1.45-1.64) | 1.41 (1.33-1.50) |
| **Ethnicity** | | | | |
| Black | 0.80 (0.78-0.83) | 0.80 (0.78-0.83) | 1.70 (1.56-1.86) | 1.26 (1.13-1.41) |
| Other and mixed | 0.72 (0.69-0.76) | 0.75 (0.72-0.79) | 1.08 (0.94-1.24) | 0.94 (0.80-1.11) |
| South Asian | 0.80 (0.78-0.82) | 0.84 (0.82-0.87) | 1.17 (1.08-1.28) | 1.07 (0.97-1.19) |
| White | 1.00 | 1.00 | 1.00 | 1.00 |
| **Region** | | | | |
| London | 0.90 (0.89-0.91) | 0.87 (0.86-0.88) | 1.26 (1.20-1.32) | 1.14 (1.08-1.20) |
| Other | 1.00 | 1.00 | 1.00 | 1.00 |
| **Urban Rural** | | | | |
| Rural | 0.88 (0.87-0.89) | 0.90 (0.89-0.91) | 0.82 (0.78-0.86) | 0.83 (0.79-0.88) |
| Urban | 1.00 | 1.00 | 1.00 | 1.00 |
| **HEALTH BEHAVIOURS / INDICATORS** | | | | |
| **Body Mass Index** | | | | |
| <18.5 (Underweight) | 4.26 (4.18-4.34) | 3.31 (3.25-3.37) | 3.99 (3.69-4.32) | 3.48 (3.24-3.73) |
| 18.5-<25 (Normal weight) | 1.00 | 1.00 | 1.00 | 1.00 |
| 25-<30 (Overweight) | 0.61 (0.60-0.62) | 0.73 (0.72-0.74) | 0.64 (0.61-0.68) | 0.76 (0.71-0.81) |
| 30-<35 (Obesity class I) | 0.65 (0.64-0.66) | 0.77 (0.76-0.79) | 0.72 (0.68-0.77) | 0.87 (0.82-0.93) |
| >=35 (Obesity class II plus) | 0.95 (0.93-0.96) | 1.11 (1.09-1.13) | 1.05 (0.98-1.13) | 1.29 (1.20-1.39) |
| **Smoking status** | | | | |
| Current smoker | 2.52 (2.48-2.56) | 2.14 (2.11-2.18) | 2.01 (1.88-2.15) | 1.91 (1.78-2.06) |
| Ex-smoker | 1.42 (1.40-1.44) | 1.26 (1.24-1.28) | 1.35 (1.28-1.43) | 1.26 (1.19-1.33) |
| Non-smoker | 1.00 | 1.00 | 1.00 | 1.00 |
| **MORBIDITY** | | | | |
| **Autoimmune condition** | | | | |
| Lupus erythematosus | 1.67 (1.52-1.83) | 1.63 (1.54-1.72) | 1.50 (1.03-2.19) | 1.14 (0.89-1.46) |
| Psoriasis | 1.14 (1.12-1.16) | 1.14 (1.12-1.16) | 1.17 (1.10-1.26) | 1.20 (1.11-1.28) |
| Rheumatoid arthritis | 1.46 (1.42-1.50) | 1.51 (1.48-1.55) | 1.55 (1.40-1.72) | 1.54 (1.42-1.67) |
| **Cardiovascular disease** | | | | |
| Cerebrovascular disease | 1.94 (1.92-1.97) | 2.06 (2.03-2.09) | 2.04 (1.94-2.15) | 2.23 (2.11-2.36) |
| Chronic heart disease | 1.91 (1.89-1.93) | 2.14 (2.11-2.17) | 1.89 (1.80-1.97) | 2.24 (2.11-2.38) |
| Hypertension | 1.30 (1.28-1.31) | 1.25 (1.23-1.26) | 1.41 (1.35-1.47) | 1.34 (1.27-1.41) |
| Venous thromboembolism | 2.28 (2.24-2.31) | 2.54 (2.50-2.59) | 2.19 (2.05-2.34) | 2.56 (2.36-2.77) |
| **Chronic respiratory disease** | | | | |
| Asthma | 1.09 (1.07-1.10) | 1.12 (1.10-1.13) | 1.06 (1.00-1.13) | 1.14 (1.08-1.21) |
| Other | 2.54 (2.50-2.57) | 2.72 (2.67-2.76) | 2.25 (2.12-2.39) | 2.50 (2.33-2.68) |
| **Neurological conditions** | | | | |
| Dementia | 3.31 (3.27-3.36) | 3.62 (3.56-3.68) | 4.75 (4.50-5.01) | 5.31 (4.99-5.64) |
| Learning disabilities | 3.32 (3.18-3.46) | 3.83 (3.66-4.01) | 4.58 (4.00-5.24) | 5.64 (4.85-6.56) |
| Other associated with respiratory infections | 2.41 (2.37-2.45) | 2.34 (2.30-2.39) | 2.60 (2.44-2.77) | 2.49 (2.31-2.67) |
| **Other comorbidity** | | | | |
| Cancer (diagnosed in last year) | 11.8 (11.4-12.1) | 13.4 (13.0-13.7) | 8.66 (7.69-9.76) | 10.4 (9.3-11.6) |
| Chronic kidney disease | 2.11 (2.09-2.14) | 1.97 (1.94-2.00) | 2.29 (2.17-2.41) | 2.20 (2.08-2.34) |
| Diabetes | 1.61 (1.59-1.63) | 1.70 (1.68-1.73) | 1.89 (1.80-1.98) | 1.98 (1.87-2.10) |
| Multimorbidity | 2.55 (2.51-2.59) | 2.66 (2.61-2.71) | 2.50 (2.35-2.66) | 2.81 (2.61-3.03) |

#### (C) By database

| | Pre-pandemic | | During Wave 1 | |
|---|---|---|---|---|
| | CPRD GOLD | CPRD Aurum | CPRD GOLD | CPRD Aurum |
| **DEMOGRAPHICS** | | | | |
| **Age** | | | | |
| 5-year increase in age | 1.66 (1.66-1.67) | 1.68 (1.68-1.69) | 1.67 (1.65-1.69) | 1.72 (1.70-1.73) |
| **Sex** | | | | |
| Female | 1.00 | 1.00 | 1.00 | 1.00 |
| Male | 1.33 (1.32-1.35) | 1.37 (1.35-1.38) | 1.35 (1.28-1.42) | 1.42 (1.37-1.48) |
| **Carstairs deprivation index quintile** | | | | |
| 1 (least deprived) | 1.00 | 1.00 | 1.00 | 1.00 |
| 2 | 1.07 (1.04-1.10) | 1.04 (1.03-1.05) | 1.02 (0.93-1.13) | 1.04 (0.99-1.10) |
| 3 | 1.23 (1.21-1.26) | 1.13 (1.12-1.15) | 1.23 (1.13-1.34) | 1.12 (1.07-1.18) |
| 4 | 1.30 (1.27-1.33) | 1.26 (1.25-1.28) | 1.25 (1.15-1.36) | 1.26 (1.20-1.32) |
| 5 (most deprived) | 1.38 (1.35-1.41) | 1.36 (1.35-1.38) | 1.43 (1.31-1.56) | 1.48 (1.41-1.56) |
| **Ethnicity** | | | | |
| Black | 0.61 (0.53-0.69) | 0.83 (0.81-0.85) | 0.98 (0.67-1.43) | 1.54 (1.43-1.65) |
| Other and mixed | 0.66 (0.60-0.72) | 0.75 (0.73-0.78) | 0.76 (0.54-1.07) | 1.06 (0.95-1.18) |
| South Asian | 0.66 (0.61-0.72) | 0.84 (0.82-0.86) | 0.84 (0.63-1.11) | 1.16 (1.09-1.25) |
| White | 1.00 | 1.00 | 1.00 | 1.00 |
| **Region** | | | | |
| London | 0.70 (0.68-0.73) | 0.92 (0.91-0.92) | 0.78 (0.68-0.88) | 1.25 (1.20-1.30) |
| Other | 1.00 | 1.00 | 1.00 | 1.00 |
| **Urban Rural** | | | | |
| Rural | 0.88 (0.86-0.89) | 0.89 (0.89-0.90) | 0.82 (0.77-0.88) | 0.83 (0.79-0.86) |
| Urban | 1.00 | 1.00 | 1.00 | 1.00 |
| **HEALTH BEHAVIOURS / INDICATORS** | | | | |
| **Body Mass Index** | | | | |
| <18.5 (Underweight) | 3.28 (3.20-3.36) | 3.61 (3.56-3.67) | 3.20 (2.90-3.53) | 3.63 (3.42-3.86) |
| 18.5-<25 (Normal weight) | 1.00 | 1.00 | 1.00 | 1.00 |
| 25-<30 (Overweight) | 0.65 (0.64-0.67) | 0.67 (0.66-0.67) | 0.66 (0.61-0.70) | 0.71 (0.68-0.74) |
| 30-<35 (Obesity class I) | 0.67 (0.66-0.69) | 0.72 (0.71-0.73) | 0.73 (0.67-0.79) | 0.81 (0.77-0.85) |
| >=35 (Obesity class II plus) | 0.96 (0.93-0.98) | 1.05 (1.03-1.06) | 1.01 (0.92-1.11) | 1.22 (1.15-1.29) |
| **Smoking status** | | | | |
| Current smoker | 2.52 (2.48-2.57) | 2.23 (2.20-2.26) | 2.30 (2.12-2.50) | 1.83 (1.73-1.94) |
| Ex-smoker | 1.42 (1.40-1.44) | 1.30 (1.28-1.31) | 1.35 (1.27-1.43) | 1.27 (1.22-1.33) |
| Non-smoker | 1.00 | 1.00 | 1.00 | 1.00 |
| **MORBIDITY** | | | | |
| **Autoimmune condition** | | | | |
| Lupus erythematosus | 1.78 (1.62-1.95) | 1.59 (1.51-1.68) | 1.73 (1.19-2.53) | 1.08 (0.84-1.38) |
| Psoriasis | 1.19 (1.16-1.22) | 1.13 (1.11-1.14) | 1.24 (1.12-1.36) | 1.18 (1.11-1.24) |
| Rheumatoid arthritis | 1.50 (1.45-1.55) | 1.50 (1.47-1.53) | 1.52 (1.32-1.74) | 1.54 (1.44-1.66) |
| **Cardiovascular disease** | | | | |
| Cerebrovascular disease | 1.94 (1.91-1.97) | 2.00 (1.97-2.02) | 2.00 (1.88-2.13) | 2.14 (2.05-2.24) |
| Chronic heart disease | 1.90 (1.87-1.92) | 2.02 (2.00-2.04) | 1.87 (1.77-1.98) | 2.04 (1.96-2.13) |
| Hypertension | 1.37 (1.35-1.39) | 1.24 (1.23-1.25) | 1.34 (1.26-1.42) | 1.38 (1.33-1.43) |
| Venous thromboembolism | 2.14 (2.10-2.19) | 2.36 (2.32-2.39) | 2.11 (1.95-2.28) | 2.29 (2.17-2.42) |
| **Chronic respiratory disease** | | | | |
| Asthma | 1.04 (1.02-1.07) | 1.12 (1.11-1.14) | 0.98 (0.90-1.07) | 1.13 (1.08-1.18) |
| Other | 2.47 (2.44-2.51) | 2.59 (2.56-2.62) | 2.30 (2.15-2.45) | 2.32 (2.21-2.44) |
| **Neurological conditions** | | | | |
| Dementia | 3.26 (3.21-3.32) | 3.45 (3.41-3.49) | 4.40 (4.14-4.69) | 5.02 (4.81-5.25) |
| Learning disabilities | 3.40 (3.20-3.63) | 3.57 (3.45-3.70) | 4.85 (3.92-5.99) | 5.08 (4.53-5.69) |
| Other associated with respiratory infections | 2.32 (2.26-2.38) | 2.40 (2.36-2.43) | 2.67 (2.42-2.94) | 2.52 (2.39-2.66) |
| **Other comorbidity** | | | | |
| Cancer (diagnosed in last year) | 9.70 (9.50-9.90) | 11.9 (11.7-12.1) | 8.20 (7.50-8.97) | 8.68 (7.99-9.44) |
| Chronic kidney disease | 2.14 (2.11-2.17) | 1.96 (1.94-1.98) | 2.18 (2.06-2.30) | 2.20 (2.10-2.29) |
| Diabetes | 1.64 (1.62-1.67) | 1.64 (1.62-1.65) | 1.81 (1.71-1.92) | 1.93 (1.86-2.01) |
| Multimorbidity | 2.33 (2.29-2.36) | 2.79 (2.75-2.83) | 2.36 (2.23-2.51) | 2.90 (2.74-3.06) |

##### (D) With and without linked data

| | Pre-pandemic | | During Wave 1 | |
|---|---|---|---|---|
| | 40 to 69 | 70 or older | 40 to 69 | 70 or older |
| **DEMOGRAPHICS** | | | | |
| **Age** | | | | |
| 5-year increase in age | 1.67 (1.67-1.68) | 1.70 (1.69-1.70) | 1.70 (1.69-1.71) | 1.76 (1.74-1.78) |
| **Sex** | | | | |
| Female | 1.00 | 1.00 | 1.00 | 1.00 |
| Male | 1.36 (1.35-1.37) | 1.36 (1.34-1.37) | 1.42 (1.37-1.47) | 1.37 (1.32-1.42) |
| **Carstairs deprivation index quintile** | | | | |
| 1 (least deprived) | 1.00 | 1.00 | 1.00 | 1.00 |
| 2 | 1.04 (1.03-1.05) | 1.04 (1.03-1.05) | 1.05 (1.00-1.10) | 1.03 (0.98-1.09) |
| 3 | 1.16 (1.14-1.17) | 1.14 (1.12-1.15) | 1.15 (1.10-1.20) | 1.12 (1.07-1.18) |
| 4 | 1.28 (1.27-1.29) | 1.27 (1.25-1.28) | 1.26 (1.20-1.31) | 1.26 (1.20-1.33) |
| 5 (most deprived) | 1.37 (1.35-1.38) | 1.36 (1.34-1.37) | 1.48 (1.42-1.55) | 1.49 (1.42-1.56) |
| **Ethnicity** | | | | |
| Black | 0.80 (0.78-0.82) | 0.76 (0.74-0.78) | 1.50 (1.40-1.61) | 1.48 (1.38-1.59) |
| Other and mixed | 0.74 (0.71-0.76) | 0.71 (0.69-0.74) | 1.02 (0.92-1.13) | 1.05 (0.94-1.16) |
| South Asian | 0.82 (0.80-0.83) | 0.74 (0.72-0.75) | 1.13 (1.06-1.21) | 1.12 (1.05-1.19) |
| White | 1.00 | 1.00 | 1.00 | 1.00 |
| **Region** | | | | |
| London | 0.89 (0.88-0.89) | 0.89 (0.88-0.90) | 1.20 (1.16-1.25) | 1.23 (1.18-1.28) |
| Other | 1.00 | 1.00 | 1.00 | 1.00 |
| **Urban Rural** | | | | |
| Rural | 0.89 (0.88-0.90) | 0.90 (0.89-0.91) | 0.82 (0.79-0.86) | 0.84 (0.81-0.88) |
| Urban | 1.00 | 1.00 | 1.00 | 1.00 |
| **HEALTH BEHAVIOURS / INDICATORS** | | | | |
| **Body Mass Index** | | | | |
| <18.5 (Underweight) | 3.67 (3.62-3.72) | 3.61 (3.56-3.66) | 3.67 (3.47-3.89) | 3.60 (3.38-3.82) |
| 18.5-<25 (Normal weight) | 1.00 | 1.00 | 1.00 | 1.00 |
| 25-<30 (Overweight) | 0.67 (0.66-0.67) | 0.67 (0.66-0.68) | 0.70 (0.67-0.73) | 0.71 (0.67-0.74) |
| 30-<35 (Obesity class I) | 0.71 (0.70-0.72) | 0.72 (0.71-0.73) | 0.80 (0.76-0.84) | 0.83 (0.78-0.87) |
| >=35 (Obesity class II plus) | 1.03 (1.02-1.05) | 1.06 (1.04-1.07) | 1.18 (1.12-1.25) | 1.24 (1.17-1.31) |
| **Smoking status** | | | | |
| Current smoker | 2.33 (2.30-2.36) | 2.23 (2.20-2.25) | 1.98 (1.88-2.09) | 1.69 (1.60-1.79) |
| Ex-smoker | 1.33 (1.32-1.35) | 1.44 (1.42-1.45) | 1.31 (1.25-1.36) | 1.30 (1.25-1.36) |
| Non-smoker | 1.00 | 1.00 | 1.00 | 1.00 |
| **MORBIDITY** | | | | |
| **Autoimmune condition** | | | | |
| Lupus erythematosus | 1.64 (1.56-1.72) | 1.62 (1.54-1.71) | 1.22 (0.99-1.50) | 1.13 (0.88-1.45) |
| Psoriasis | 1.14 (1.13-1.16) | 1.13 (1.11-1.15) | 1.19 (1.13-1.25) | 1.17 (1.10-1.24) |
| Rheumatoid arthritis | 1.50 (1.48-1.53) | 1.50 (1.48-1.53) | 1.55 (1.45-1.65) | 1.56 (1.45-1.68) |
| **Cardiovascular disease** | | | | |
| Cerebrovascular disease | 2.02 (2.00-2.04) | 1.99 (1.97-2.01) | 2.15 (2.07-2.24) | 2.19 (2.09-2.28) |
| Chronic heart disease | 2.03 (2.01-2.05) | 2.00 (1.98-2.02) | 2.05 (1.98-2.13) | 2.08 (2.00-2.17) |
| Hypertension | 1.28 (1.27-1.29) | 1.23 (1.21-1.24) | 1.38 (1.33-1.43) | 1.41 (1.36-1.46) |
| Venous thromboembolism | 2.43 (2.39-2.46) | 2.36 (2.33-2.39) | 2.38 (2.26-2.51) | 2.33 (2.20-2.46) |
| **Chronic respiratory disease** | | | | |
| Asthma | 1.11 (1.10-1.12) | 1.12 (1.11-1.13) | 1.11 (1.06-1.15) | 1.14 (1.09-1.19) |
| Other | 2.63 (2.60-2.66) | 2.59 (2.56-2.63) | 2.38 (2.27-2.49) | 2.36 (2.24-2.48) |
| **Neurological conditions** | | | | |
| Dementia | 3.48 (3.44-3.51) | 3.44 (3.40-3.48) | 5.02 (4.82-5.23) | 5.12 (4.90-5.34) |
| Learning disabilities | 3.54 (3.43-3.65) | 3.85 (3.73-3.98) | 5.04 (4.56-5.58) | 5.49 (4.93-6.10) |
| Other associated with respiratory infections | 2.39 (2.36-2.42) | 2.39 (2.35-2.42) | 2.57 (2.45-2.70) | 2.58 (2.45-2.73) |
| **Other comorbidity** | | | | |
| Cancer (diagnosed in last year) | 12.6 (12.4-12.8) | 15.1 (14.8-15.4) | 9.52 (8.76-10.34) | 10.8 (10.0-11.8) |
| Chronic kidney disease | 2.06 (2.03-2.08) | 1.95 (1.93-1.97) | 2.27 (2.18-2.36) | 2.24 (2.15-2.35) |
| Diabetes | 1.66 (1.64-1.68) | 1.75 (1.73-1.76) | 1.95 (1.88-2.02) | 2.11 (2.02-2.20) |
| Multimorbidity | 2.62 (2.59-2.65) | 5.12 (5.05-5.19) | 2.67 (2.54-2.80) | 5.00 (4.71-5.31) |

#### (E) Ethnicity in London versus the rest of the UK

| | Before Wave 1 | | | During Wave 1 | | |
|---|---|---|---|---|---|---|
| | Overall | London only | Other | Overall | London only | Other |
| White | 1.00 | 1.00 | 1.00 | 1.00 | 1.00 | 1.00 |
| South Asian | 0.82 (0.80-0.83) | 0.81 (0.79-0.84) | 0.84 (0.82-0.86) | 1.13 (1.06-1.21) | 1.10 (1.00-1.22) | 1.03 (0.93-1.13) |
| Black | 0.80 (0.78-0.82) | 0.84 (0.82-0.87) | 0.77 (0.73-0.80) | 1.50 (1.40-1.61) | 1.39 (1.27-1.52) | 1.33 (1.17-1.52) |
| Other and mixed | 0.74 (0.71-0.76) | 0.71 (0.67-0.75) | 0.78 (0.75-0.81) | 1.02 (0.92-1.13) | 1.06 (0.92-1.23) | 0.86 (0.74-1.01) |
| Missing | 1.13 (1.12-1.14) | 0.96 (0.93-0.99) | 1.13 (1.12-1.14) | 1.04 (1.01-1.08) | 0.97 (0.87-1.08) | 1.05 (1.02-1.09) |

#### (F) Missing data

| | Before Wave 1 | During Wave 1 |
|---|---|---|
| Ethnicity | | |
| Black | 0.80 (0.78-0.82) | 1.50 (1.40-1.61) |
| Other and mixed | 0.74 (0.71-0.76) | 1.02 (0.92-1.13) |
| South Asian | 0.82 (0.80-0.83) | 1.13 (1.06-1.21) |
| White | 1.00 | 1.00 |
| Missing | 1.13 (1.12-1.14) | 1.04 (1.01-1.08) |
| Body Mass Index | | |
| <18.5 (Underweight) | 3.67 (3.62-3.72) | 3.67 (3.47-3.89) |
| 18.5-<25 (Normal weight) | 1.00 | 1.00 |
| 25-<30 (Overweight) | 0.67 (0.66-0.67) | 0.70 (0.67-0.73) |
| 30-<35 (Obesity class I) | 0.71 (0.70-0.72) | 0.80 (0.76-0.84) |
| 35-<40 (Obesity class II) | 1.03 (1.02-1.05) | 1.18 (1.12-1.25) |
| Missing | 1.05 (1.03-1.06) | 1.05 (0.99-1.11) |
| Smoking status | | |
| Current smoker | 2.33 (2.30-2.36) | 1.98 (1.88-2.09) |
| Ex-smoker | 1.33 (1.32-1.35) | 1.31 (1.25-1.36) |
| Non-smoker | 1.00 | 1.00 |
| Missing | 0.87 (0.85-0.88) | 1.19 (1.10-1.29) |
